# Leveraging Global Genetics Resources to Enhance Polygenic Prediction Across Ancestrally Diverse Populations

**DOI:** 10.1101/2025.03.27.25324773

**Authors:** Oliver Pain

## Abstract

**Introduction:** Genome-wide association studies (GWAS) from multiple ancestral populations are increasingly available, offering opportunities to improve the accuracy and equity of polygenic scores (PGS). Several methods now aim to leverage multiple GWAS sources, but predictive performance and computational efficiency across contexts remain unclear, especially in the absence of individual-level tuning data.

**Methods:** This study evaluates a comprehensive set of PGS methods across African (AFR), East Asian (EAS), and European (EUR) ancestry groups for 10 complex traits, using summary statistics from the Ugandan Genome Resource, Biobank Japan, and UK Biobank. Single-source PGS were derived using methods including DBSLMM, lassosum, LDpred2, MegaPRS, pT+clump, PRS-CS, QuickPRS, and SBayesRC. Multi-source approaches included PRS-CSx, TL-PRS, X-Wing, and combinations of independently optimised single-source scores. A key contribution is the introduction of a novel application of the LEOPARD method to estimate optimal linear combinations of population-specific PGS using only summary statistics. All analyses were implemented using the GenoPred software pipeline.

**Results:** In AFR and EAS populations, PGS combining ancestry-aligned and European GWAS outperformed single-source models. Linear combinations of independently optimised scores consistently outperformed current jointly optimised multi-source methods, while being substantially more computationally efficient. The LEOPARD extension offered a practical solution for tuning these combinations when only summary statistics were available, achieving performance comparable to tuning with individual-level data.

**Conclusion:** These findings highlight a flexible and generalisable framework for multi-source PGS construction. The GenoPred pipeline enables researchers to tailor methods to data availability and study goals, supporting more equitable, accurate, and accessible polygenic prediction.

## Introduction

Genome-wide association studies (GWAS) have identified thousands of genetic variants associated with a range of outcomes (Abdellaoui et al., 2023; Cerezo et al., 2025). Polygenic scores (PGS) harness this wealth of data by aggregating the effects of numerous genetic variants to estimate an individual’s genetic predisposition to specific traits or diseases (Choi et al., 2020). As GWAS sample sizes continue to grow and novel PGS methods emerge, the predictive power and utility of PGS in research and clinical settings have significantly improved. Of particular interest is the use of PGS for disease risk stratification (Fuat et al., 2024; Khera et al., 2018), which creates opportunities to enhance disease prevention, early diagnosis, and targeted treatment (Kullo et al., 2022; Lewis & Vassos, 2020; Wray et al., 2021).

However, a major limitation of current GWAS is the overrepresentation of individuals of European (EUR) ancestry, which leads to a substantial decrease in PGS performance across populations (Duncan et al., 2019; Martin et al., 2019; Privé et al., 2022; Wang et al., 2022). PGS derived from EUR-based GWAS typically perform better in EUR populations than in non-EUR populations, such as those of African (AFR) or East Asian (EAS) ancestry. This disparity reduces the clinical utility of PGS, and their current use would exacerbate health inequalities. The increasing availability of GWAS within non-EUR populations has prompted the development of PGS methodology that can leverage multiple population-specific GWAS (Kachuri et al., 2024), aiming to improve predictive performance and ensure broader applicability in diverse populations.

PGS are typically calculated by aggregating genetic variants across the genome, weighting each variant according to how strongly it relates to a specific trait. Various PGS methods exist for estimating these variant weights based on GWAS results. PGS methods can be broadly categorised as *single-source* or *multi-source* (Figure 1). Single-source PGS methods, such as PRS-CS, derive the variant weights from a single GWAS (Ge et al., 2019). Multi-source methods, in contrast, incorporate multiple GWAS from different populations to improve prediction. *Independently optimised* multi-source methods apply single-source PGS methods separately to each GWAS, producing multiple ancestry-specific PGS that are later linearly combined to optimise prediction for a given target population. These are typically denoted by appending “-multi” to the base method (e.g., PRS-CS-multi). More recently, *jointly optimised* multi-source methods have been developed, such as PRS-CSx, which leverage cross-population data to enhance effect size estimation and improve robustness (Miao et al., 2023; Ruan et al., 2022; Zhao et al., 2022). Like independently optimised methods, they produce separate PGS for each population, which can also be linearly combined to refine prediction.

**Figure 1.**
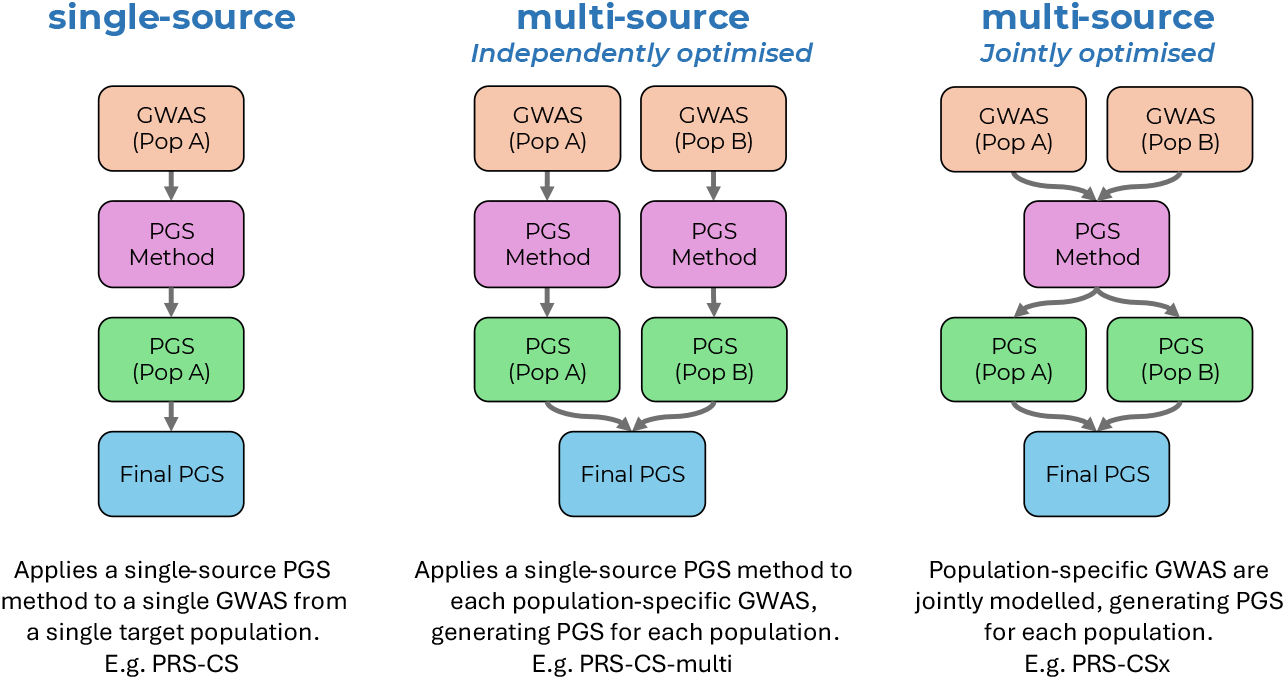
Overview of PGS methods leveraging GWAS from one or more populations. Pop = Population.

Previous research has shown that integrating multiple GWAS from different populations enhances PGS accuracy, but the optimal approach remains uncertain. Several studies suggest that jointly optimised PGS methods outperform independently optimised approaches (Gunn et al., 2025; Miao et al., 2023; Ruan et al., 2022). However, one recent study indicates that independently optimised methods can surpass currently available jointly optimised strategies (Zheng et al., 2024). This is likely because advances in single-source methodologies have not yet been incorporated into jointly optimised multi-source methods. Additionally, independently optimised methods are reported to require fewer computational resources (Zheng et al., 2024), making them more practical for large-scale applications. A comprehensive and independent evaluation of currently available multi-source PGS strategies is needed to determine the most effective approach.

An important consideration when comparing PGS methods is whether they can be tuned using only GWAS summary statistics. This approach, often termed “pseudovalidation” or “auto” modelling, allows tuning of the final PGS without needing individual-level genotype or phenotype data. Most PGS methods generate multiple scores by varying hyperparameters, such as *p*-value thresholds or shrinkage parameters. Multi-source methods generate separate PGS for each ancestry-specific GWAS. To improve prediction in a given target population, these scores can be combined linearly to find an optimal weighted sum. Determining the best weights, however, requires an additional tuning step. While individual-level data can be used for this tuning, it is often inaccessible. Alternative techniques (such as sample-splitting to avoid overfitting) introduce additional complexity. In contrast, approaches that rely solely on GWAS summary statistics for tuning offer a more practical solution. However, while a range of summary statistic tuning methods exist for optimising individual PGS hyperparameters, fewer approaches are available for tuning the optimal linear combination of population-specific PGS scores. Notably, no summary-statistic approach has been tested for independently optimised multi-source methods. One promising solution is the LEOPARD method, which estimates the optimal linear combination of population-specific PGS without requiring individual-level data. Currently, LEOPARD is used only within the X-Wing multi-source PGS method and has not yet been explored for other PGS methods (Miao et al., 2023). Extending this approach to other multi-source methods could provide a generalisable strategy for tuning the optimal linear combination of population-specific PGS. Addressing this gap is essential for enhancing the robustness and applicability of multi-source PGS approaches.

The GenoPred pipeline is an easy-to-use, high-performance, reference-standardised, and reproducible workflow for polygenic scoring (Pain et al., 2024). The GenoPred reference-standardised framework has been used in previous studies for comparing polygenic scoring methods (Pain et al., 2021). The GenoPred pipeline makes the identified leading approaches more accessible by facilitating their implementation. Although earlier versions of GenoPred supported GWAS and target samples from various ancestral populations, they were limited to single-source PGS methods, restricting the pipeline’s ability to fully leverage the increasingly available ancestrally diverse GWAS data.

In this study, we use the GenoPred pipeline to systematically evaluate the performance of various PGS methods and modelling approaches across African (AFR), East Asian (EAS), and European (EUR) populations. Furthermore, we introduce a novel application of the LEOPARD method as a generalisable and efficient solution for estimating the optimal linear combination of population-specific PGS. The aim is to optimise and broaden access to multi-source PGS methods that leverage ancestrally diverse datasets, contributing to more equitable, accurate, and accessible polygenic prediction.

## Methods

### Overview

This study evaluated the performance of PGS methods and modelling approaches across diverse populations using GWAS and target datasets of EUR, EAS, and AFR ancestry (Figure 2). EUR GWAS summary statistics were generated within a training subset of European ancestry individuals in the UK Biobank sample (UKB)(Bycroft et al., 2018). Publicly available EAS and AFR GWAS were obtained from Biobank Japan (BBJ) (Sakaue et al., 2021) and the Ugandan Genome Resource (UGR) (Gurdasani et al., 2019), respectively. An independent target sample of EUR, EAS and AFR ancestry individuals within UKB were used to evaluate the predictive performance of the PGS. To compare PGS methods, 10 traits available in UKB, BBJ, and UGR were selected to represent a range of genetic architectures.

**Figure 2.**
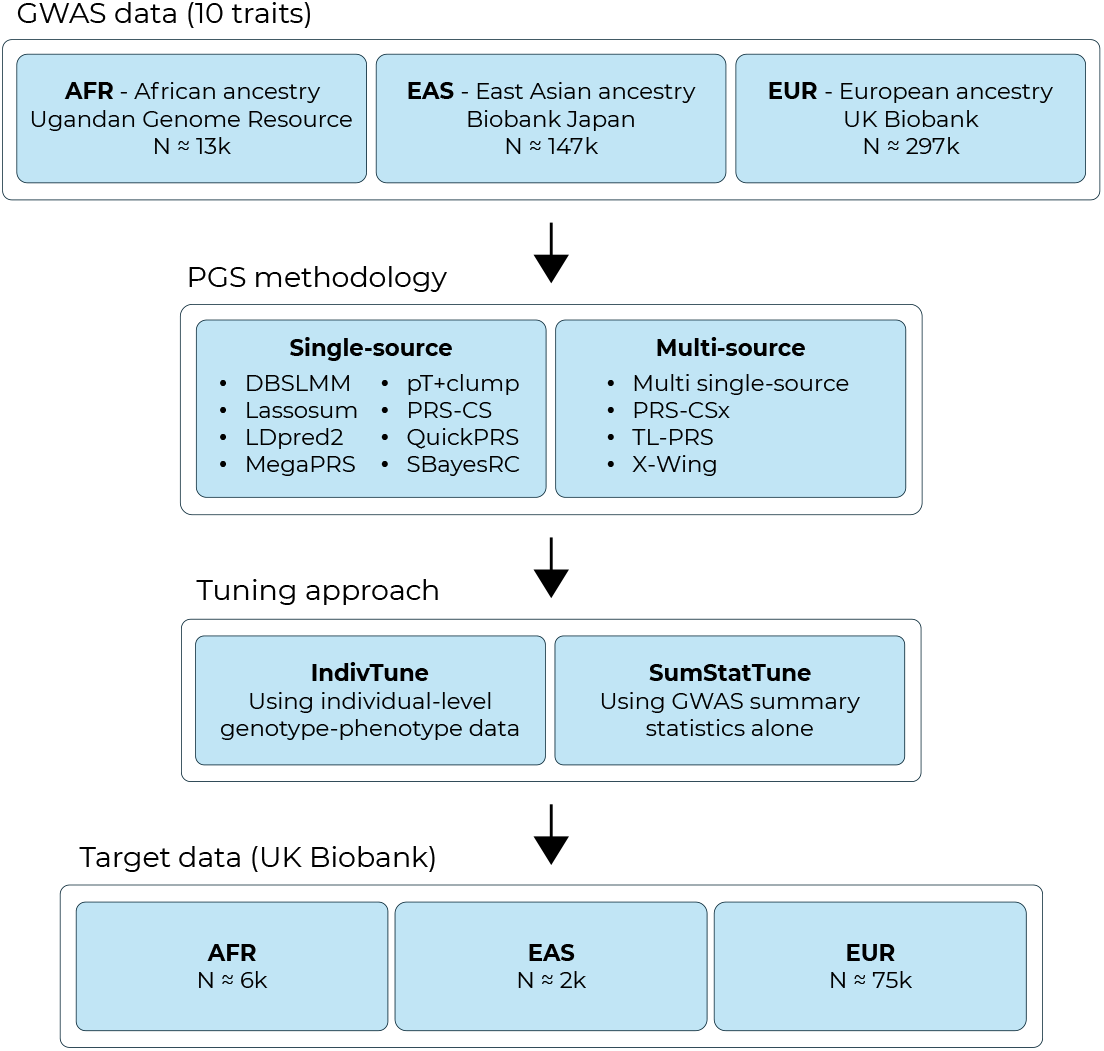
Study design overview. ‘Multi single-source’ refers to the independently optimised multi-source approach of combining multiple single-source PGS. `N` indicates median sample size.

### UK Biobank

UKB is a prospective cohort study that recruited >500,000 individuals aged between 40–69 years across the United Kingdom (Bycroft et al., 2018). The UKB received ethical approval from the North West – Haydock Research Ethics Committee (reference 16/NW/0274).

UKB was used to generate GWAS summary statistics for EUR ancestry and to evaluate the predictive utility of PGS in individuals of EUR, EAS, and AFR ancestry. To avoid sample overlap between the EUR GWAS and EUR target samples, EUR individuals in UKB were split into a training subset for GWAS (80%), and testing subset for evaluating PGS (20%).

#### Ancestry inference

The ancestry of UKB individuals was genetically inferred, matching individuals to populations within the reference genetic dataset, a combination of samples from 1000 Genomes phase 3 and the Human Genome Diversity Project (1KG+HGDP) (1000 Genomes Project Consortium, 2015; Bergström et al., 2020). Outlier individuals were then removed from each inferred population. Ancestry inference and outlier detection was performed using the GenoPred pipeline (Pain et al., 2024), using the imputed UKB genetic data as input, in PLINK2 binary format (.pgen) (Chang et al., 2015), and filtered to include variants with a minor allele frequency of ≥1% and an imputation INFO score > 0.4.

In brief, GenoPred estimates each individual’s probability of belonging to a reference population using a multinomial elastic net model trained on six genetic principal components derived from the reference dataset. These components are projected onto UKB, and individuals are assigned to a population if the predicted probability is >0.95. For outlier detection, GenoPred uses principal components analysis within each inferred UKB population to capture population-specific structure and batch effects, identifying outliers based on k-means clustering centroids. Full details are available in the GenoPred documentation (Pain et al., 2024).

#### Outcome trait preparation

Outcome trait data were extracted using the ukbkings R package (Hanscombe, 2022). Unrelated individuals were identified via the UKB-provided kinship matrix and the GreedyRelated software v1.2.1 (see URLs). These individuals were then split into their inferred ancestral populations. EUR individuals were further split into training (80%) and testing subsets (20%). Within each group, the outcome was inverse rank-based normalised, and then covariates were regressed out (Pain et al., 2018). Covariates included age, sex, and the first 20 within-UKB genetic principal components (PCs). The residuals were then scaled and centred to have a mean of 0 and standard deviation of 1.

### GWAS summary statistics

GWAS summary statistics for EUR, EAS, and AFR populations were obtained from UKB, BBJ, and UGR respectively (Gurdasani et al., 2019; Sakaue et al., 2021). Publicly available BBJ and UGR GWAS summary statistics were downloaded for this study. GWAS summary statistics for EUR were derived in the training subset of UKB for this study using PLINK2 (Chang et al., 2015).

Data for 28 metabolic, cardiovascular, and anthropometric traits were available in UKB, BBJ and UGR. Descriptive statistics for these traits are provided in Table S1 (including BBJ/UGR download links and UKB field codes).

Quality control of GWAS summary statistics was performed using the GenoPred pipeline. In brief: strand-ambiguous variants are removed; variants are aligned to the 1KG+HGDP reference (restricted to HapMap3 variants; see URLs); and results were filtered based on imputation quality (INFO < 0.9), minor allele frequency (MAF < 0.01 or MAF discrepancies with reference > 0.2), valid p-values (0 < P ≤ 1), presence of unique SNP IDs, and acceptable sample size ranges. Missing BETA coefficients and standard errors (SE) are calculated if absent. Full details are available in the GenoPred documentation (Pain et al., 2024).

### Selecting traits for comparison

To minimise computational costs associated with running all PGS methods repeatedly, a subset of 10 traits was selected for downstream analyses. These traits were chosen to represent a range of genetic architectures, including SNP-based heritability (SNP-*h*^2^) and polygenicity. LD score regression (LDSC) with GWAS summary statistics was used to estimate SNP-*h*^2^ (Bulik-Sullivan et al., 2015). The AVENGEME software (Additive Variance Explained and Number of Genetic Effects Method of Estimation) was used to estimate SNP-*h*^2^ and the proportion of variants with no effect on the trait (pi0), a metric representing the inverse of polygenicity (Palla & Dudbridge, 2015). AVENGEME uses PGS associations across a range of *p*-value thresholds to estimate these parameters. The GenoPred pipeline was used to calculate the PGS in UKB using the *p*-value thresholding and clumping (*p*T+clump) method. Association analysis was then conducted in R version 4.2.3 (R Core Team, 2025).

Traits were initially filtered to retain those with a positive SNP-*h*^2^ point estimate from both LDSC and AVENGEME in EUR, EAS, and AFR populations. The SNP-*h*^2^ and pi0 estimates from AVENGEME (using EUR data) were then used to randomly select 10 traits that capture a range of these values, and therefore a range of genetic architectures. EUR AVENGEME results were used as the estimates were most accurate due to the larger sample size compared to EAS and AFR populations. The selected traits are indicated in Table S1.

### PGS methodology

A range of leading summary statistic PGS methods, including single- and multi-source, were applied using the GenoPred pipeline. Single-source methods include DBSLMM (Yang & Zhou, 2020), lassosum (Mak et al., 2017), LDpred2 (Privé et al., 2020), MegaPRS (Zhang et al., 2021), pT+clump (Chang et al., 2015), PRS-CS (Ge et al., 2019), QuickPRS (Fast variant MegaPRS, see URLs), and SBayesRC (Zheng et al., 2024). Each of these methods were also evaluated as independently optimised multi-source methods. This involves applying the single-source PGS method to each GWAS and linearly combining the population-specific PGS to optimise prediction in a given target population. Jointly optimised multi-source methods include PRS-CSx (Ruan et al., 2022), and X-Wing (Miao et al., 2023). In contrast to these, the TL-PRS method represents a distinct multi-source approach: it starts with an existing “baseline” PGS model (a set of SNP weights tuned in one population) and further tunes it using GWAS data from the target population’s ancestry. TL-PRS is a post-hoc adjustment applied to an already-developed PGS, rather than a method that simultaneously derives weights from multiple GWAS.

These methods were selected based on their performance in previous literature. The multi-source method BridgePRS (Hoggart et al., 2024) was not evaluated in this study because it currently cannot produce score files without access to individual-level phenotype and genotype data. Although several other PGS methods require individual-level data to tune the final PGS, they produce a series or score files without the need for individual-level training data, in keeping with the reference-standardised framework of the GenoPred pipeline.

As noted, the TL-PRS method is distinct from other multi-source methods; therefore, TL-PRS was evaluated separately to other methods. The originally proposed workflow for TL-PRS involves two tuning steps: first tuning a baseline PGS model to select hyperparameters, and then further tuning that model using the target population’s GWAS to select the learning rate (gradient). For simplicity, in our evaluation we used baseline PGS models that were tuned using summary statistics alone. This meant that individual-level data were required only for the second step of tuning the baseline PGS with the target population’s GWAS. When GWAS summary statistics are available for two populations, TL-PRS can be run in both directions (a procedure referred to as MTL-PRS). In other words, one can tune a PGS from Population A using Population B’s GWAS, and vice versa, then linearly combine the two population-specific PGS. PGS adjusted using TL-PRS are denoted by appending ‘TL-’ to the method’s name (e.g. TL-SBayesRC and TL-SBayesRC-multi). A schematic representation of the TL-PRS workflow is shown in Figure S1.

The same 1KG+HGDP reference data was used for all PGS methods except PRS-CS, PRS-CSx, and X-Wing. For those three methods, we used the 1000 Genomes reference data provided with their software, as it is not straightforward to create a custom reference in the required format. As a sensitivity analysis, a collection of methods was run using only 1KG reference individuals to assess the impact of different reference data.

### Estimating optimal linear combination of population-specific PGS

As mentioned above, no summary-statistic approach has been tested for tuning the linear combination of population-specific PGS from independently optimised multi-source methods. Among jointly optimised methods, PRS-CSx applies a simple inverse-variance meta-analysis to combine population-specific PGS into a single score for any population (Ruan et al., 2022). In contrast, X-Wing employs the LEOPARD method (Miao et al., 2023), a more advanced approach that estimates the optimal linear combination of population-specific PGS for a given target population, allowing greater flexibility when certain GWAS are more relevant than others. There is no summary-statistic tuning approach for TL-PRS.

This study presents a novel application of X-Wing’s LEOPARD method: we use LEOPARD to estimate the optimal linear combination of population-specific PGS for independently optimised multi-source approaches. LEOPARD involves splitting GWAS into training and testing subsets, training the PGS weights in the training subset, evaluating the PGS in the testing subset, and from this estimate the appropriate weight of population-specific PGS for a given target population. Given that single-source PGS methods showed broadly similar performance, we used QuickPRS within the LEOPARD analysis to generate the PGS weights (Figure 3). This assumes that the optimal weights for combining population-specific PGS would not depend strongly on which single-source method was used, and thus QuickPRS would be representative of the others. The PGS weights from LEOPARD were scaled to correspond to each population-specific PGS having a SD of 1, to improve their applicability to scaled PGS from other methods. Note that LEOPARD estimates the optimal linear combination of one PGS from each population. In practice, this means we apply the LEOPARD-derived weights to one PGS per population (specifically, the PGS that was produced by each single-source method’s summary-statistic-only tuning approach).

**Figure 3.**
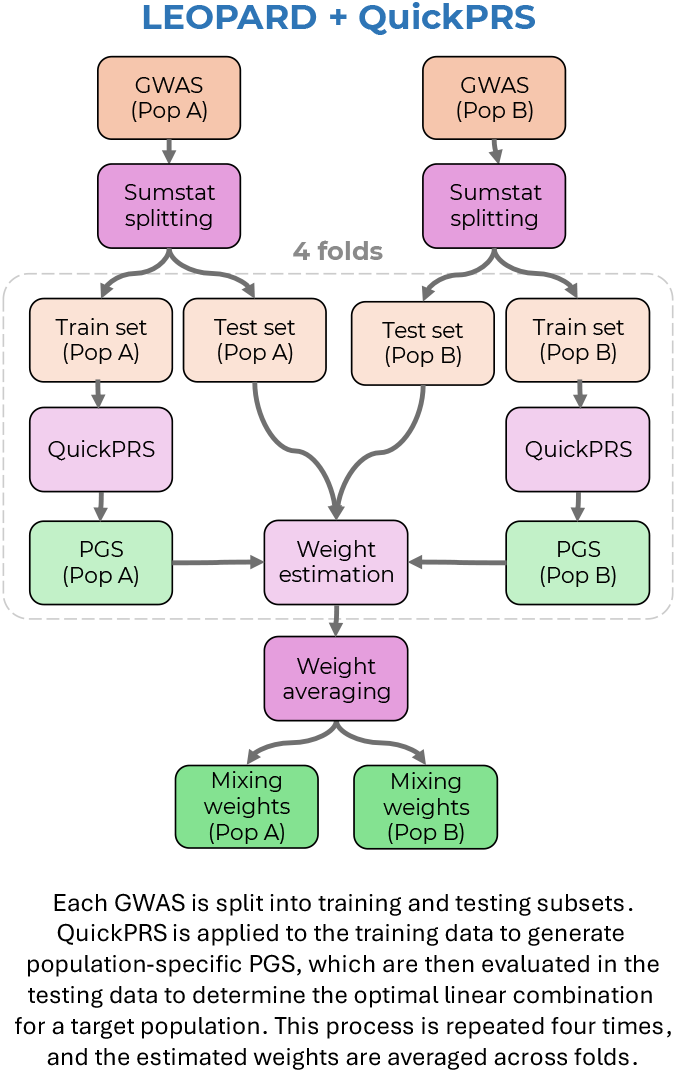
Overview of LEOPARD + QuickPRS approach. Pop = Population.

### Evaluating PGS

The predictive utility of the PGS was assessed using 10-fold cross-validation. The training subset was used to identify the optimal hyperparameters for each population-specific PGS and to determine the optimal linear combination of these population-specific PGS. The final PGS model was then evaluated in the held-out test subset. PGS models were evaluated using R, with model evaluation performed using the model_builder_top1.R script within the GenoPred repository (see URLs).

To assess whether the best PGS method varied across traits, an “All” model was also derived. The “All” model considered every PGS method: for single-source approaches it selected the best-performing population-specific PGS, and for multi-source approaches it identified the optimal linear combination of population-specific PGS. This approach provides a robust benchmark representing the highest achievable performance under both single- and multi-source scenarios, against which other PGS methods can be compared.

### Evaluating prediction accuracy

Prediction accuracy was evaluated as the Pearson correlation between the observed and predicted outcome values. To compare PGS methods and modelling approaches, the correlations between observed and predicted values of each model were statistically compared using the Hotelling-Williams test (Steiger, 1980) as implemented by the ‘psych’ R package’s ‘paired.r’ function, with the correlation between model predictions of each method specified to account for their non-independence. A two-sided test was used when calculating p-values.

The correlations between predicted and observed values were then pooled across all traits for each PGS method. These correlations (and their variances) were aggregated using the “BHHR” method (Cooper et al., 2019), as implemented in the ‘MAd’ R package’s ‘agg’ function. An outcome trait correlation matrix was used in this meta-analytic aggregation to account for the non-independence of the traits within each target population.

### Computational benchmark

GenoPred is a Snakemake pipeline, so the runtime and peak memory of each PGS method were recorded using the Snakemake benchmark functionality (Mölder et al., 2021). These benchmarks reflect the time and memory used by each PGS method as implemented in the GenoPred pipeline, and they should not deviate substantially from the resource usage observed in other implementations of the same methods. All analyses were performed using the King’s Computational Research, Engineering and Technology Environment (CREATE) (King’s College London, 2024).

## Results

This study applied 10 PGS methods to GWAS for 10 traits from AFR, EAS, and EUR populations, and evaluated the PGS in independent AFR, EAS and EUR target samples from the UKB. The predictive utility and computational efficiency of PGS methods were compared when tuned using either individual-level data (IndivTune) or summary-statistics alone (SumStatTune) (Figure 2).

### Trait Selection and Descriptives

Heritability and polygenicity estimates for all 28 available traits (in AFR, EAS, and EUR populations) are provided in Table S1. The 10 traits selected (covering a range of SNP-based heritability and polygenicity) were: body mass index, body weight, haemoglobin, HDL-cholesterol, height, mean corpuscular haemoglobin concentration, neutrophil count, platelet count, systolic blood pressure, and total cholesterol. SNP-based heritability estimates (from EUR GWAS) ranged roughly from 5% to 24%, and polygenicity ranged from about 3% to 15%. The median GWAS sample sizes were approximately 13k for AFR, 147k for EAS, and 297k for EUR. The median target sample sizes were approximately 6k for AFR, 2k for EAS, and 75k for EUR. Across traits, the variance explained by the pT+clump PGS (using ancestry-aligned GWAS) ranged from 0.17% to 4.20% in AFR, 0.66% to 9.06% in EAS, and 1.88% to 17.89% in EUR (Table S2). These results confirm that the study design — comprising the GWAS, target data, and traits selected — had sufficient information to capture polygenic prediction, and that the traits are diverse in genetic architecture.

### Single-Source PGS methods

The predictive performance of single-source PGS in AFR and EAS target samples was first evaluated, using either ancestry-aligned or EUR GWAS for training (Figure 4). Across all traits, PGS trained on EUR GWAS outperformed those trained on AFR GWAS in AFR target samples. This suggests that the larger EUR GWAS sample sizes provided greater predictive power despite ancestry mismatch. In EAS target samples, PGS trained on EAS GWAS performed similarly to those trained on EUR GWAS. This indicates that the ancestral relevance of the EAS GWAS compensated for its smaller sample size, yielding prediction accuracy comparable to the larger EUR GWAS. As expected, the absolute predictive performance was lower in the AFR target than the EAS target. This likely reflects the smaller sample size of the AFR GWAS and the greater genetic distance between the AFR population and the EUR population (Figure S2).

**Figure 4.**
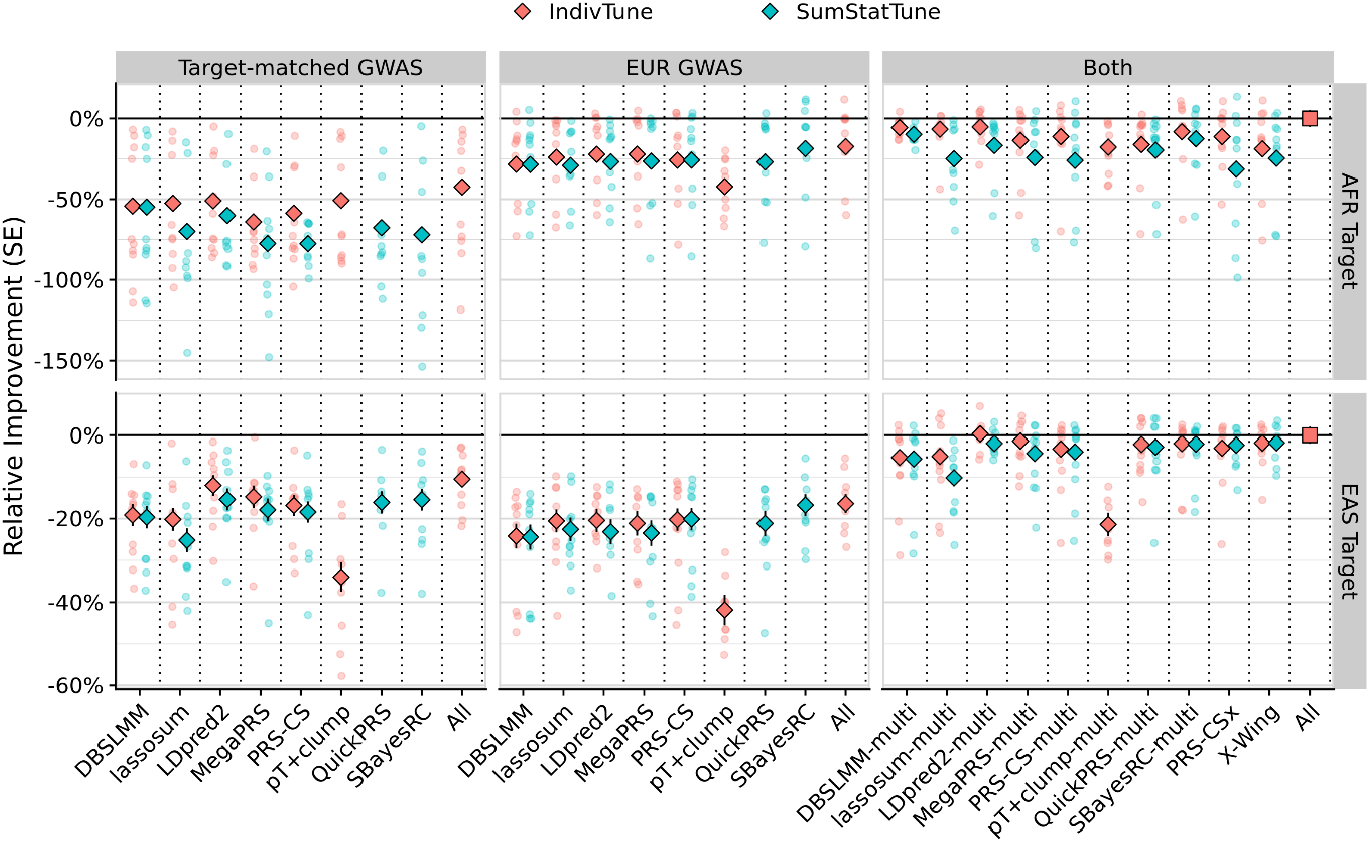
Relative predictive performance of PGS methods in AFR and EAS target populations. The y-axis shows the relative improvement in predictive performance compared to the multi-source ‘All’ model, with error bars representing the standard error. The diamond-shaped points indicate the average difference across traits, with small circular points indicating trait-specific differences. Colours indicate whether PGS methods were trained using individual-level data (IndivTune) or GWAS summary statistics alone (SumStatTune). Facet columns represent the source of the GWAS data used for PGS derivation, including target ancestry-aligned (‘Target-matched’) GWAS, European (EUR) GWAS, and combined target ancestry-aligned and EUR GWAS (‘Both’). Facet rows show performance in African (AFR) and East Asian (EAS) target populations. The ‘All’ model was derived by selecting the best-performing population-specific PGS across all methods for single-source approaches and identifying the optimal linear combination of population-specific PGS for multi-source methods. This ensures that the ‘All’ model provides a robust benchmark, reflecting the highest achievable predictive utility under both single- and multi-source scenarios.

The relative performance of single-source PGS methods was broadly consistent regardless of which population’s GWAS was used (Figure 4), though two notable exceptions emerged. First, the simple pT+clump method performed relatively well when using AFR GWAS compared to EAS and EUR GWAS. Second, the performance of SumStatTune approaches for MegaPRS, QuickPRS, and SBayesRC performed worse when using AFR GWAS compared to EAS or EUR GWAS.

Models selecting the best PGS across all methods yielded performance gains (relative to the single best method) of about 17% in AFR targets (p = 2×10^−3^) and 1.7% in EAS targets (p = 0.34) (Figure 4; Figures S3–S4; Table S3). This suggests that no single method consistently performs best across all traits with AFR GWAS. In contrast, with EAS GWAS a single method (LDpred2) performed well across most traits. When using EUR GWAS, SBayesRC performed well across all traits – A model considering PGS across all methods did not provide a statistically significant improvement over SBayesRC (only +1.5% in AFR target, p = 0.37; and +0.4% in EAS target, p = 0.83). Detailed trait-specific performance of all PGS methods is provided in Figures S5–S14 and Table S2.

In the EUR target population, the relative performance ranking of the single-source PGS methods was similar to that observed in other populations (Figure S15).

### Multi-Source PGS methods

Multi-source PGS methods demonstrated greater predictive power than single-source approaches in both AFR and EAS target populations (Figure 4). Both jointly and independently optimised multi-source methods (which combine multiple ancestry-specific PGS) generally outperformed the single-source PGS. For example, LDpred2-multi outperformed the standard single-population LDpred2 PGS by 21.8% in AFR target (p = 1×10^−28^) and 14.0% in EAS target (p = 2×10^−9^) (Table S3). The relative improvement of LDpred2-multi over LDpred2 varied across traits, ranging from 0.3% to 37.3% in AFR and 3.0% to 22.2% in EAS (Table S2).

On average, the performance was similar across the different multi-source methods. LDpred2-multi performed best when individual-level tuning data were available (IndivTune), whereas SBayesRC-multi was the most effective when tuning PGS using summary statistics alone (SumStatTune). Models selecting the best PGS across all multi-source methods yielded an additional improvement of 5.6% in AFR (p = 1×10^-3^), but showed no significant improvement in EAS (-0.3%, p = 0.82) (Figure 4).

The jointly optimised methods (PRS-CSx and X-Wing) did not outperform independently optimised approaches. On average, PRS-CSx showed no improvement over its independently optimised counterpart (PRS-CS-multi) in either AFR or EAS targets. X-Wing performed worst with AFR data but with EAS data it achieved results comparable to the best methods. A sensitivity analysis using only 1KG reference individuals (as opposed to 1KG+HGDP) showed no change in performance for any of the methods (Figure S16).

As expected, using TL-PRS to tune a EUR-based PGS with a target-matched GWAS significantly improved prediction accuracy compared to the unadjusted EUR PGS. However, applying TL-PRS to independently optimised multi-source PGS resulted in only marginal improvements (Figure S17). For example, MTL-SBayesRC-multi — which applies TL-PRS to the SBayesRC-multi scores — improved prediction accuracy by only 1.2% in AFR (p = 0.19) and 2.3% in EAS (p = 0.02), relative to SBayesRC-multi without TL adjustment.

Multi-source PGS provided a small but statistically significant improvement over using only a EUR PGS (Figures S15, S22–S23, Table S3). For example, on average SBayesRC-multi improved prediction accuracy over single-population SBayesRC by 0.1% (p = 8×10^−4^) when using EUR+AFR data, and by 0.9% (p = 5×10^−33^) when using EUR+EAS data.

### Impact of tuning PGS without individual-level data

Tuning PGS with individual-level data (IndivTune) generally improved prediction accuracy compared to tuning based on GWAS summary statistics alone (SumStatTune). Among single-source methods, the SumStatTune approach performed worst for certain methods when using AFR GWAS.

Among jointly optimised multi-source methods, X-Wing’s summary-statistic tuning approach (LEOPARD) performed well. PRS-CSx also performed well using SumStatTune with EAS data but performed poorly with AFR data. Notably, this study’s novel implementation of LEOPARD with QuickPRS as a SumStatTune approach for independently optimised multi-source methods also showed strong performance. For example, in the case of QuickPRS-multi, tuning the linear combination of population-specific PGS with individual-level data provided only marginal improvements: about +4.4% in AFR and +0.6% in EAS, respectively. The optimal weights for combining population-specific PGS (when estimated using individual-level data) were highly correlated across the different PGS methods (Figures S18– S21). This suggests that the LEOPARD weights derived from QuickPRS generalise well to other single-source PGS methods. Figure 5 compares the performance of models containing SumStatTune PGS, where population-specific weighting was done using either LEOPARD or individual-level data.

**Figure 5.**
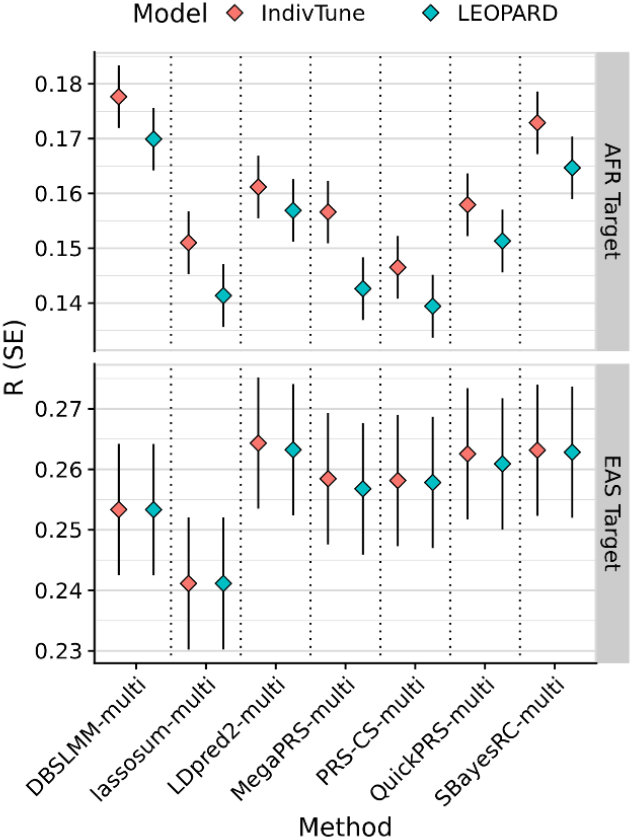
Performance of LEOPARD for weighting SumStatTune population-specific PGS. The y-axis shows the correlation (R) between predicted and observed values, with error bars representing the standard error. The x-axis lists independently optimised multi-source PGS methods, where population-specific PGS were derived using a summary-statistics-only method from the corresponding single-source method (e.g., LDpred2-auto model). Colours indicate whether population-specific PGS weights were estimated using individual-level data (IndivTune) or using the LEOPARD method with QuickPRS (LEOPARD). The top panel shows results for the African (AFR) target population, and the bottom panel shows results for the East Asian (EAS) target population.

Currently, there is no summary-statistic-based approach for tuning TL-PRS. This means TL-PRS adjustments can only be applied when individual-level data are available.

### Computational benchmark

The average time and memory required by each method are shown in Table 1. With 10 CPU (central processing unit) cores available and using the recommended tuning approach for each method, most methods complete within 30 minutes. However, some methods were substantially slower: PRS-CS, PRS-CSx, and X-Wing required roughly 4.4, 6.8, and 34.1 hours per GWAS, respectively.

**Table 1.** Computation resource required for different methods as implemented within GenoPred, using 10 cores.

| Method | Time (hrs) | Memory (GB) |
| --- | --- | --- |
| DBSLMM | 0.15 | 1.15 |
| lassosum | 0.08 | 7.30 |
| LDpred2 | 0.38 | 20.54 |
| MegaPRS | 0.85 | 12.51 |
| PRS-CS | 4.40<sup>b</sup> | 10.57 |
| pT+clump | 0.02 | 0.83 |
| QuickPRS | 0.06 | 4.87 |
| SBayesRC | 0.38 | 4.4 |
| PRS-CSx<sup>a</sup> | 6.84<sup>b</sup> | 15.18 |
| X-Wing<sup>a</sup> | 34.12<sup>c</sup> | 48.7 |
| LEOPARD<br>(QuickPRS)<sup>a</sup> | 0.23 | 8.22 |

<sup>a</sup> Time per GWAS (total time divided by 2). Memory with 2 of input GWAS.
<sup>b</sup> Time taken with grid search (4 $\phi$ parameter + auto). Auto model only will be 20% of total.
<sup>c</sup> Time taken with X-Wing with LEOPARD. Without LEOPARD, will be 20% of total.

Runtime depends strongly on the tuning procedure used. The reported times for PRS-CS and PRS-CSx assume a grid search over five different global shrinkage parameters (phi). If instead the SumStatTune (auto) approach is used, their runtimes drop to roughly 20% of those defaults — only about 0.88 and 1.37 hours, respectively. Similarly, X-Wing runs much faster if LEOPARD (the SumStatTune weighting step for population-specific PGS) is skipped, cutting its runtime to ∼6.82 hours (about 20% of the default). This speed-up occurs because the LEOPARD step requires re-estimating SNP effects four additional times per GWAS.

X-Wing, an extension of PRS-CS and PRS-CSx, is inherently slower than other PGS methods. In contrast, this studies novel application of LEOPARD with QuickPRS enables much faster estimation of population-specific PGS weights — completing in just 14 minutes per GWAS.

Thus, the independently optimised multi-source approach — using single-source PGS methods and combining them with LEOPARD+QuickPRS weights — is far more computationally efficient than jointly optimised PGS methods like PRS-CSx or X-Wing. For instance, with 10 cores and individual-level tuning, applying LDpred2 to two GWAS takes 46 minutes, whereas X-Wing takes 13.64 hours. If using a summary-statistic-only tuning approach, applying SBayesRC to two GWAS and estimating the linear combination using LEOPARD+QuickPRS takes 1.25 hours, compared to 68.24 hours for X-Wing.

With 10 cores, adjusting a single PGS model using TL-PRS took approximately 20 minutes and required 31 GB of memory.

## Discussion

This study presented a comprehensive comparison of leading PGS methods applied to datasets from multiple ancestral populations. It evaluated predictive performance and computational efficiency of both single-source and multi-source PGS methods, within and across populations, using individual-level tuning and summary statistic-only tuning approaches. As expected, multi-source methods offered significantly improved prediction over single-source methods. Notably, independently optimised multi-source methods achieved competitive prediction accuracy while being substantially more computationally efficient than the jointly optimised multi-source methods. Additionally, this study developed a novel extension of the LEOPARD method using QuickPRS, to provide a summary statistic-based approach for weighting population-specific PGS from independently optimised multi-source methods.

All the methods evaluated in this study have been integrated into the GenoPred pipeline, which is publicly available and designed for broad accessibility (see URLs). These findings, alongside prior research, underscore the importance of incorporating GWAS data from multiple populations to optimise PGS prediction in ancestrally diverse samples. Importantly, computationally efficient, summary statistic-only methods (now fully implemented in GenoPred) offer practical solutions for researchers working with limited individual-level data or resources.

This study can guide researchers in selecting appropriate PGS methods based on the availability of GWAS data, target data, and computational resources. For large EUR GWAS, SBayesRC demonstrated superior performance across all traits — notably, without needing any individual-level tuning data. SBayesRC also performed well with EAS GWAS; however, LDpred2 surpassed it when individual-level tuning data was available for EAS. The advantage of LDpred2 over SBayesRC was more pronounced when using AFR GWAS summary statistics, where several methods requiring individual-level data for tuning performed similarly to LDpred2. In general, no single method consistently outperformed all others across traits with AFR GWAS, indicating that a model tuning across PGS methods could enhance prediction accuracy.

In practise, for AFR or EAS GWAS, when individual-level training data are available, one should apply both SBayesRC and LDpred2 and select the best-performing PGS across methods. If individual-level training data are unavailable or limited, SBayesRC alone is a strong choice. When GWAS data from multiple populations are accessible, population-specific PGS can be linearly combined using either individual-level data or the LEOPARD method (which requires summary statistics only). Given the high correlation between population-specific weights across PGS methods, a computationally efficient method like QuickPRS is suitable for generating the LEOPARD weights. This strategy (implemented in the GenoPred pipeline) greatly improves efficiency without sacrificing accuracy.

This study indicates that currently available jointly optimised multi-source methods do not offer an advantage over independently optimised multi-source methods, and they come with a much higher computational cost. This conclusion contrasts with some earlier reports that jointly optimised methods outperform those independently optimised (Miao et al., 2023; Ruan et al., 2022; Wang et al., 2023; Zhao et al., 2022). However, this study’s findings are consistent with the recent SBayesRC study (Zheng et al., 2024), suggesting that incorporating functional annotations into PGS provides a more significant benefit than joint optimisation alone. It is possible that future jointly optimised multi-source methods, especially if they incorporate such features, could surpass independently optimised approaches. For now, though, independently optimised approaches remain the recommended choice given their computational efficiency.

Although the TL-PRS post-hoc adjustment improved prediction when applied to a single-source PGS (compared to no adjustment), it showed no meaningful improvement when applied to jointly optimised multi-source PGS. Within the scenarios tested in this study, the additional computational burden and tuning complexity of TL-PRS did not justify its use for multi-source PGS.

Several limitations of this study should be acknowledged. Firstly, the reference populations were defined broadly and had limited sample sizes (especially for AFR), which may have impacted PGS performance due to linkage disequilibrium (LD) reference panel misspecification. Refining the reference panels to better match each GWAS population’s LD structure could improve certain methods’ performance. Secondly, using HapMap3 variants as the default in GenoPred might influence the relative performance of PGS methods.

Denser genome coverage could enhance methods that leverage functional annotations, such as MegaPRS, QuickPRS, and SBayesRC (Zheng et al., 2024). Thirdly, this study did not evaluate approaches for admixed target individuals. Future research could investigate scoring approaches that account for local ancestry when aggregating population-specific PGS for an individual (Hou et al., 2024; Marnetto et al., 2020). Finally, scenarios where a single GWAS includes individuals from multiple ancestries were not addressed. In such cases, it is recommended to either specify a reference population that matches the majority ancestry or generate a custom reference dataset that reflects the ancestry proportions in the GWAS.

In conclusion, as GWAS data from diverse populations become increasingly available, multi-source approaches should be prioritised to enhance prediction accuracy in ancestrally diverse target samples. The GenoPred pipeline facilitates this by providing an accessible, robust, and computationally efficient framework to apply state-of-the-art PGS methods, even in the absence of individual-level tuning data. Future methodological work should aim to integrate the strengths of single-source methods into new jointly optimised multi-source methods to maximise predictive performance. Nevertheless, despite the gains from multi-source methods, large disparities in predictive accuracy between European and non-European target populations remain. Closing this gap will require significantly expanding and improving the diversity of GWAS datasets, which is crucial for more equitable and accurate polygenic prediction moving forward.

## Code availability

GenoPred homepage: https://opain.github.io/GenoPred/

GreedyRelated: https://gitlab.com/choishingwan/GreedyRelated

QuickPRS: https://dougspeed.com/quick-prs/

model_builder_top1.R: https://github.com/opain/GenoPred/blob/gwas_grouping/Scripts/model_builder/model_builder_top1.R

## Data availability

GWAS summary statistics from BBJ and UGR were publicly available (see Table S1). The UK Biobank data was accessed via project 82087 - For access, go to: https://www.ukbiobank.ac.uk/enable-your-research/apply-for-access. HapMap3 SNP-list: https://doi.org/10.5281/zenodo.7773502.

## Acknowledgements

OP thanks Michelle Kamp and Florian Privé for feedback on the manuscript; Doug Speed, Eva Krapohl, and Remo Monti for insightful discussions; Ammar Al-Chalabi for fellowship support; and Cathryn Lewis for contributions to the broader GenoPred research programme. OP also thanks the developers of all PGS methodologies evaluated in this study.

OP is supported by a Sir Henry Wellcome Postdoctoral Fellowship [222811/Z/21/Z]. The funders had no role in study design, data collection and analysis, decision to publish, or preparation of the manuscript.

This research was conducted under UK Biobank application 82087.

