## Supplementary Figures for "Leveraging Global Genetics Resources to Enhance Polygenic Prediction Across Ancestrally Diverse Populations"

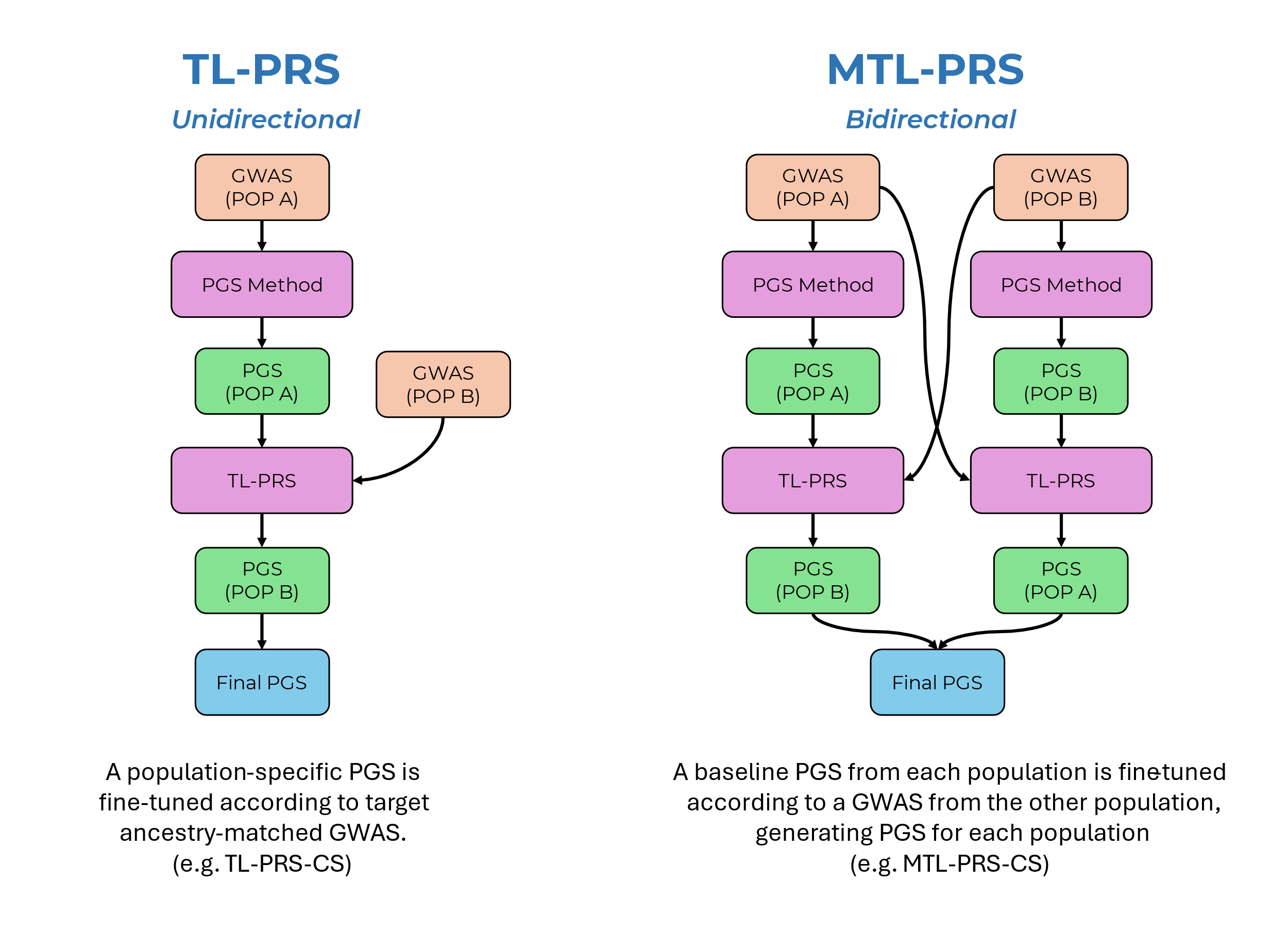

Figure S1. Schematic representation of TL-PRS method. TL-PRS involves tuning an existing PGS model from population A based on GWAS summary statistics from population B. An extension is MTL-PRS, whereby TL-PRS is used bidirectionally to tune PGS models from population A and B using GWAS summary statistics from population B and A respectively, generating two population-specific PGS that can then be linearly combined.

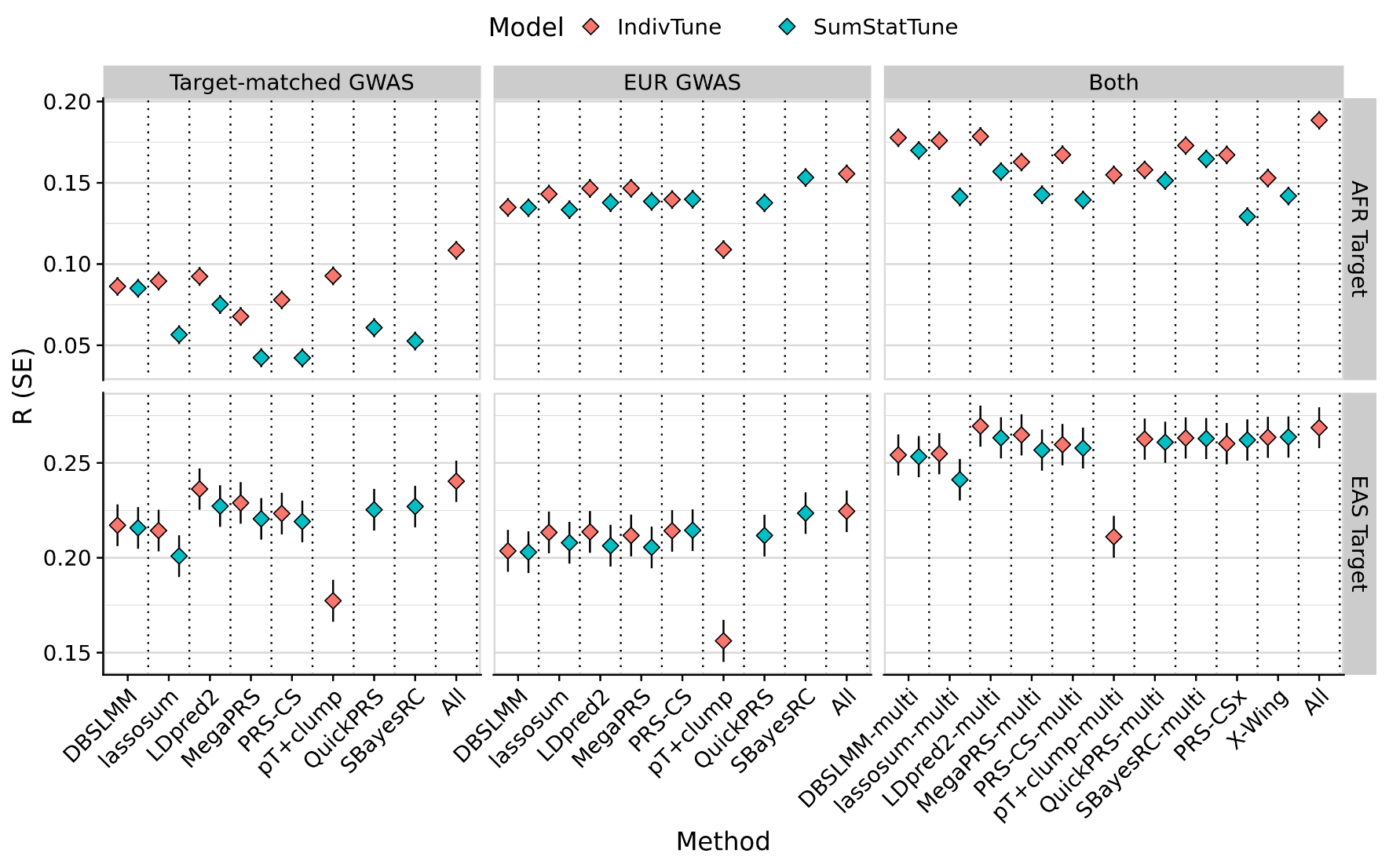

Figure S2. Average absolute predictive utility of PGS methods in AFR and EAS target populations. The y-axis indicates the average correlation between predicted and observed values across traits, with error bars showing the standard error. Colours distinguish whether PGS methods were trained using individual-level data (IndivTrain) or GWAS summary statistics alone (SumStatTrain). 'Target-matched GWAS', 'EUR GWAS', and 'Both' facets show PGS performance using target ancestry-aligned, European, or combined GWAS data, respectively. 'AFR Target' and 'EAS Target' facets show performance in AFR and EAS samples. 'All' models in the 'Both' facet represent the best population-specific PGS selected across multi-source methods. In the 'Target-matched GWAS' and 'EUR GWAS' facets, 'All' models represent the best population-specific PGS from single-source methods.

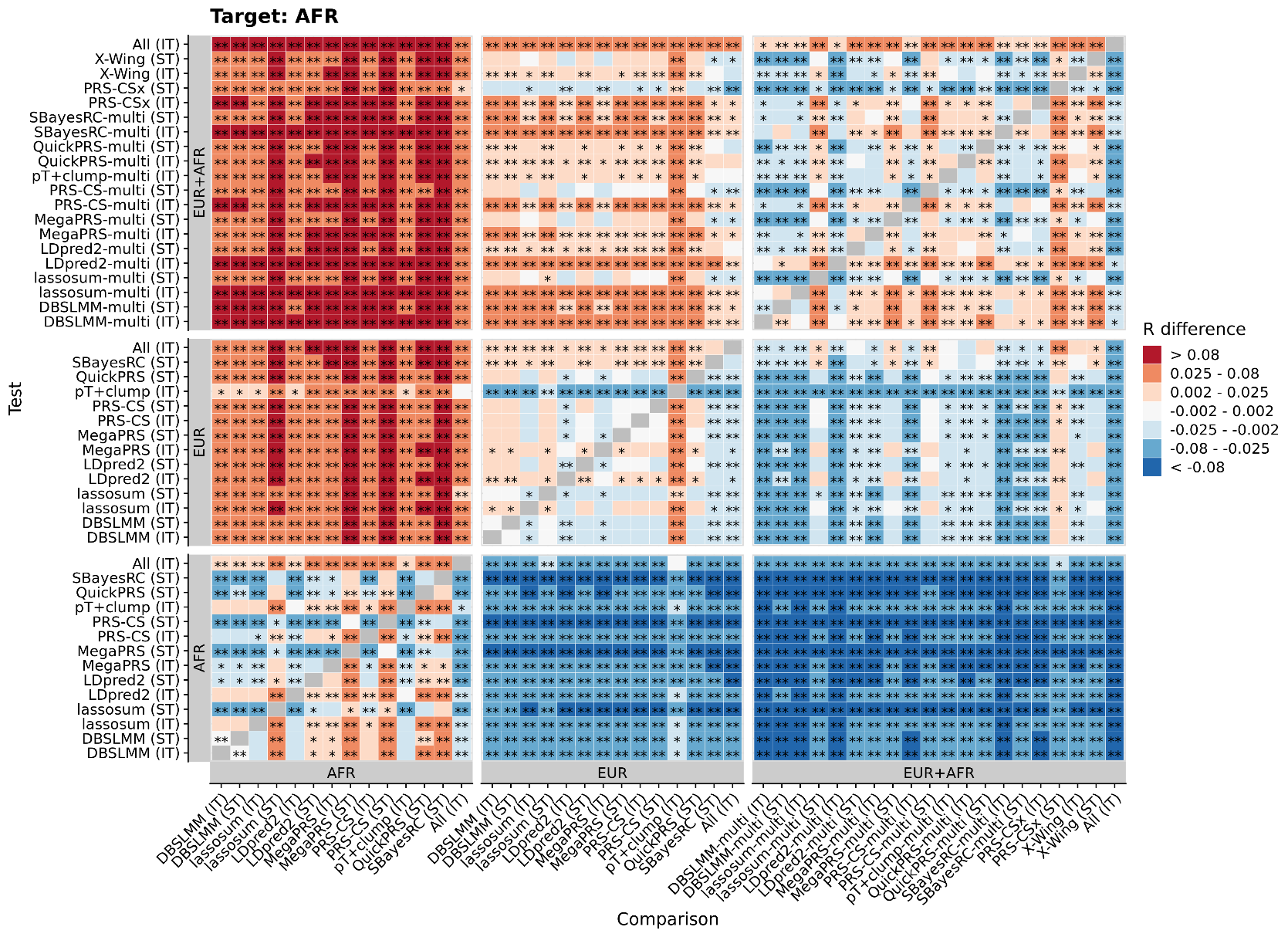

Figure S3. Pairwise comparison between all methods in AFR target sample, showing average difference in observed-expected correlation. R difference = Test correlation minus Comparison correlation. Red/orange colouring indicates the Test method (shown on Y axis) performed better than the Comparison method (shown on X axis). Shows only results based on the UKB target sample when using the 1KG reference. * = p<0.05 * = p<1×10−3. P-values are two-sided. IT = IndivTrain, PGS model tuned using individual-level data. ST = SumStatTrain, PGS model tuned using GWAS summary statistics alone.

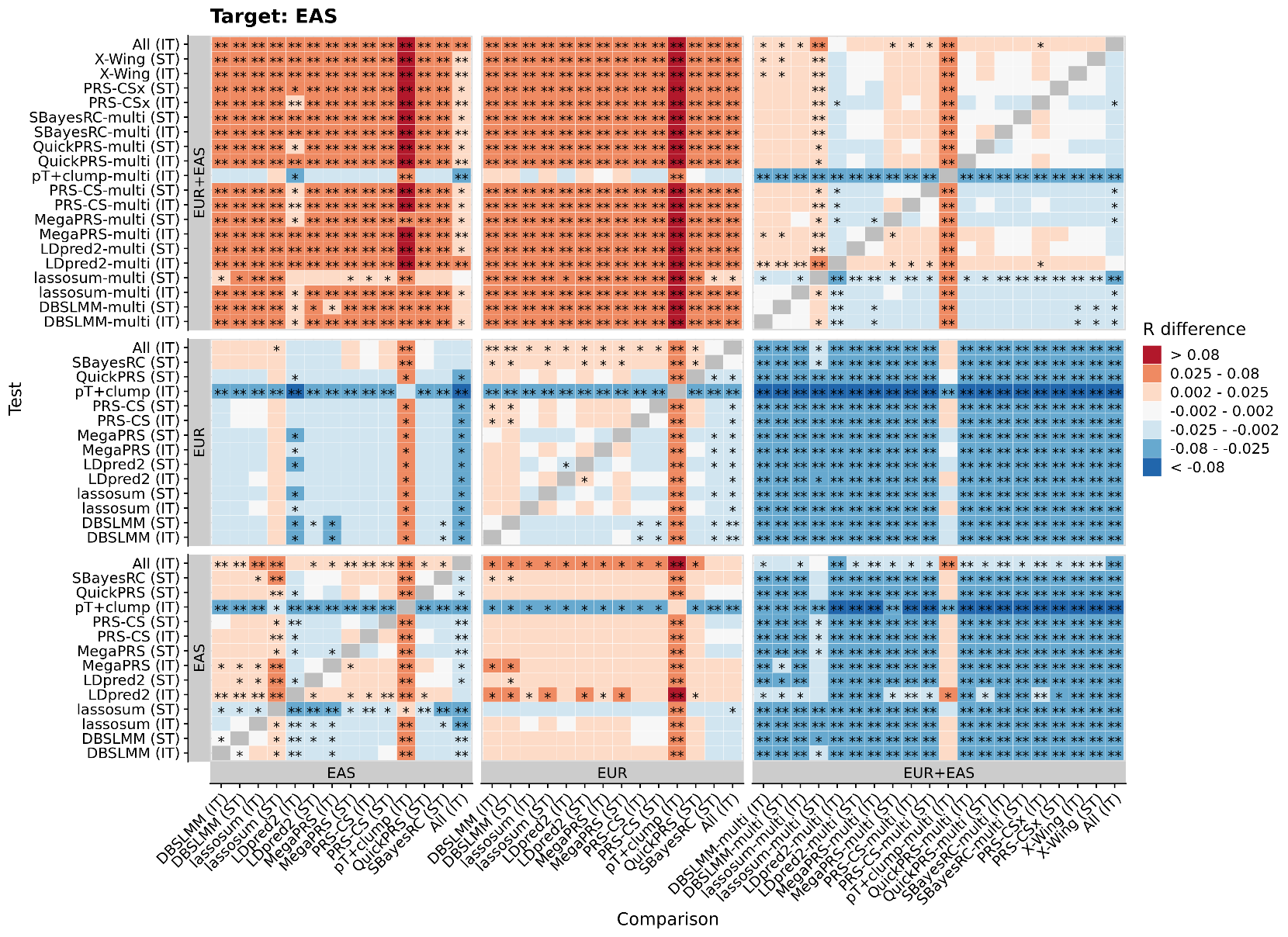

Figure S4. Pairwise comparison between all methods in EAS target sample, showing average difference in observed-expected correlation. R difference = Test correlation minus Comparison correlation. Red/orange colouring indicates the Test method (shown on Y axis) performed better than the Comparison method (shown on X axis). Shows only results based on the UKB target sample when using the 1KG reference. * = p<0.05 * = p<1×10−3. P-values are two-sided. IT = IndivTrain, PGS model tuned using individual-level data. ST = SumStatTrain, PGS model tuned using GWAS summary statistics alone.

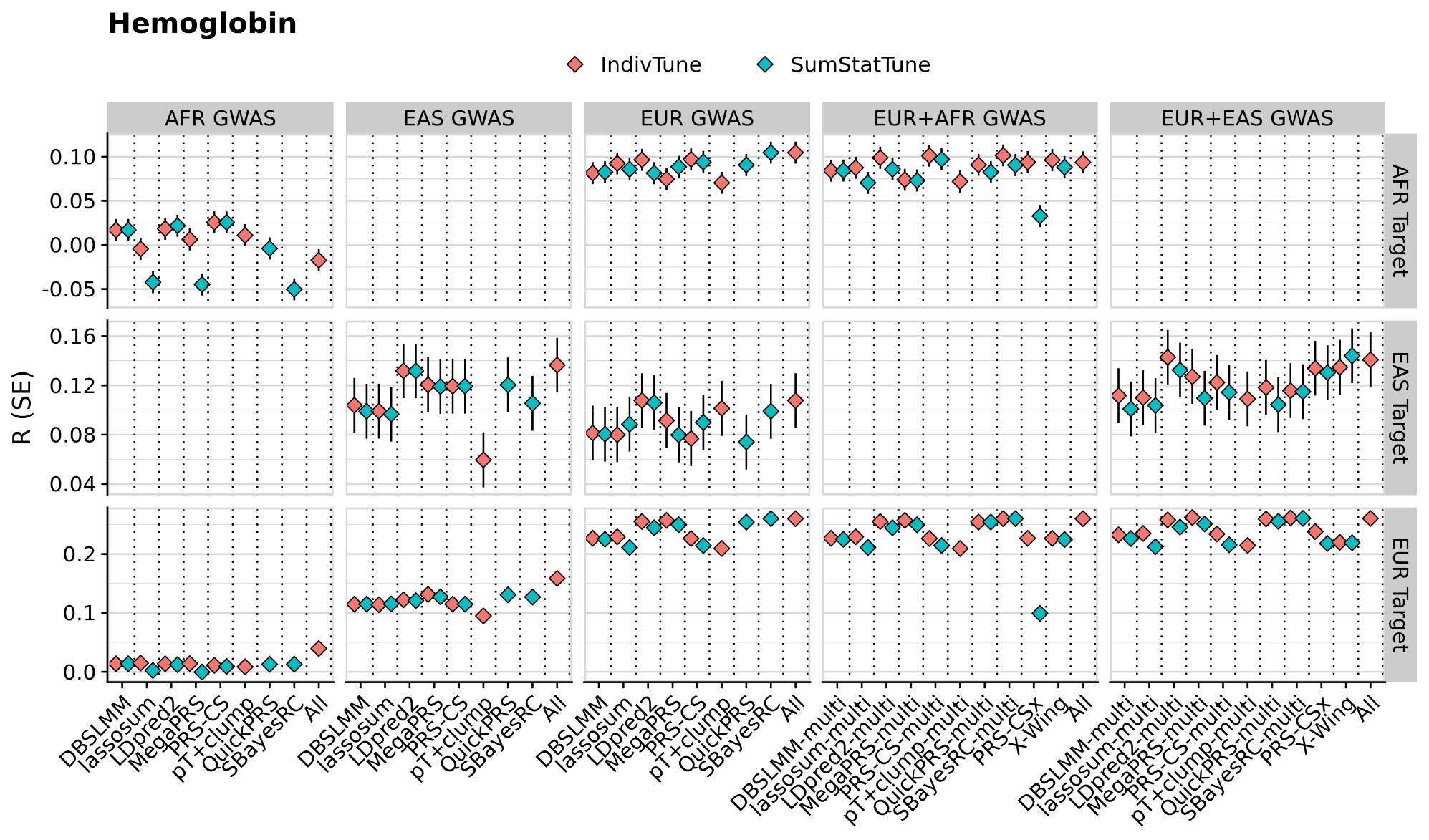

Figure S5. Predictive utility of PGS methods for Hemoglobin. The y-axis shows the correlation (R) between predicted and observed trait levels, with error bars representing the standard error. Colours differentiate between PGS methods trained using individual-level data (IndivTrain) and those trained using GWAS summary statistics (SumStatTrain). Facet columns indicate the GWAS data source used for PGS derivation, including African (AFR GWAS), East Asian (EAS GWAS), European (EUR GWAS), combined African and European (EUR+AFR GWAS), and combined East Asian and European (EUR+EAS GWAS) data. Facet rows represent performance in African (AFR Target), East Asian (EAS Target), and European (EUR Target) populations.

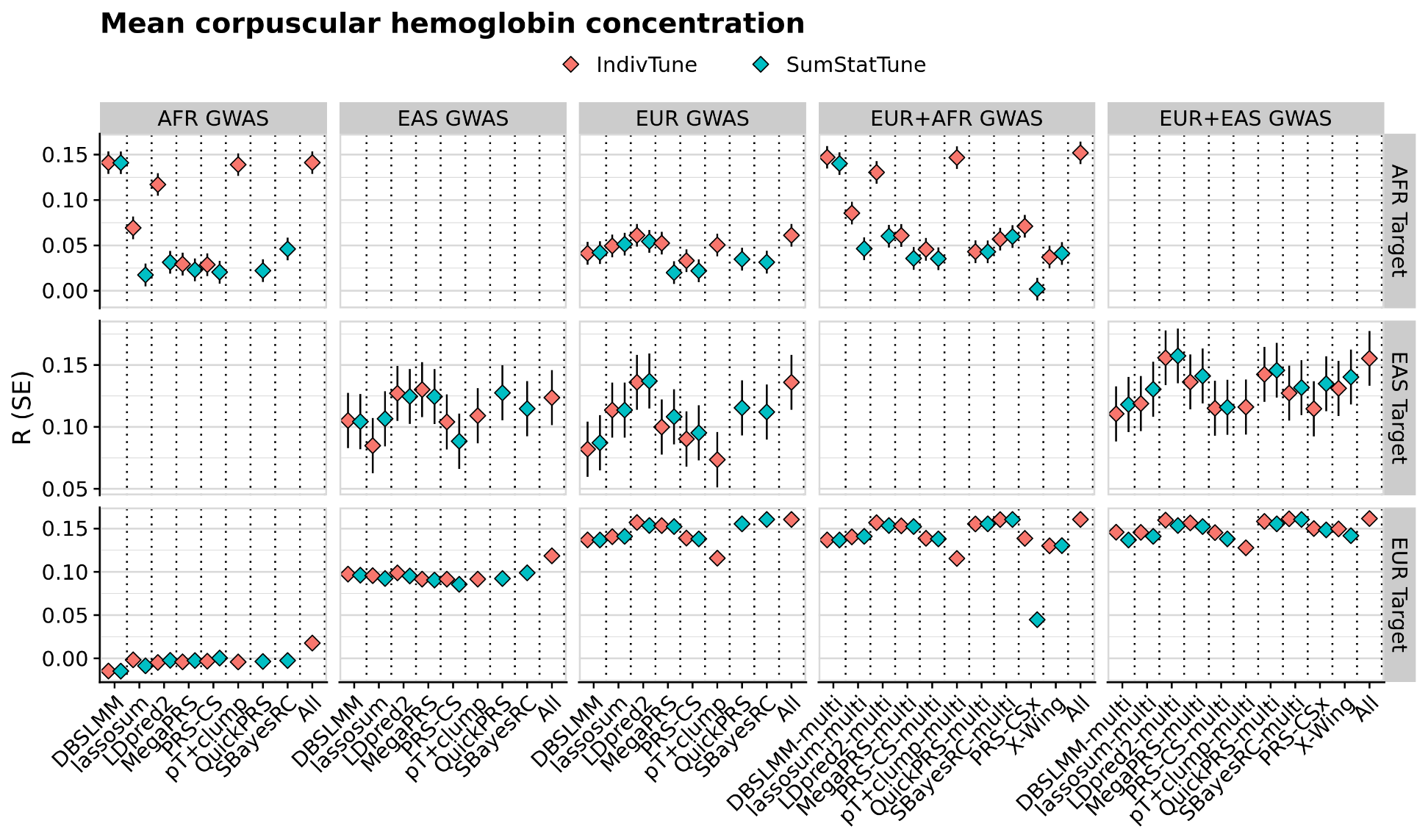

Figure S6. Predictive utility of PGS methods for Mean corpuscular hemoglobin concentration. The y-axis shows the correlation (R) between predicted and observed trait levels, with error bars representing the standard error. Colours differentiate between PGS methods trained using individual-level data (IndivTrain) and those trained using GWAS summary statistics (SumStatTrain). Facet columns indicate the GWAS data source used for PGS derivation, including African (AFR GWAS), East Asian (EAS GWAS), European (EUR GWAS), combined African and European (EUR+AFR GWAS), and combined East Asian and European (EUR+EAS GWAS) data. Facet rows represent performance in African (AFR Target), East Asian (EAS Target), and European (EUR Target) populations.

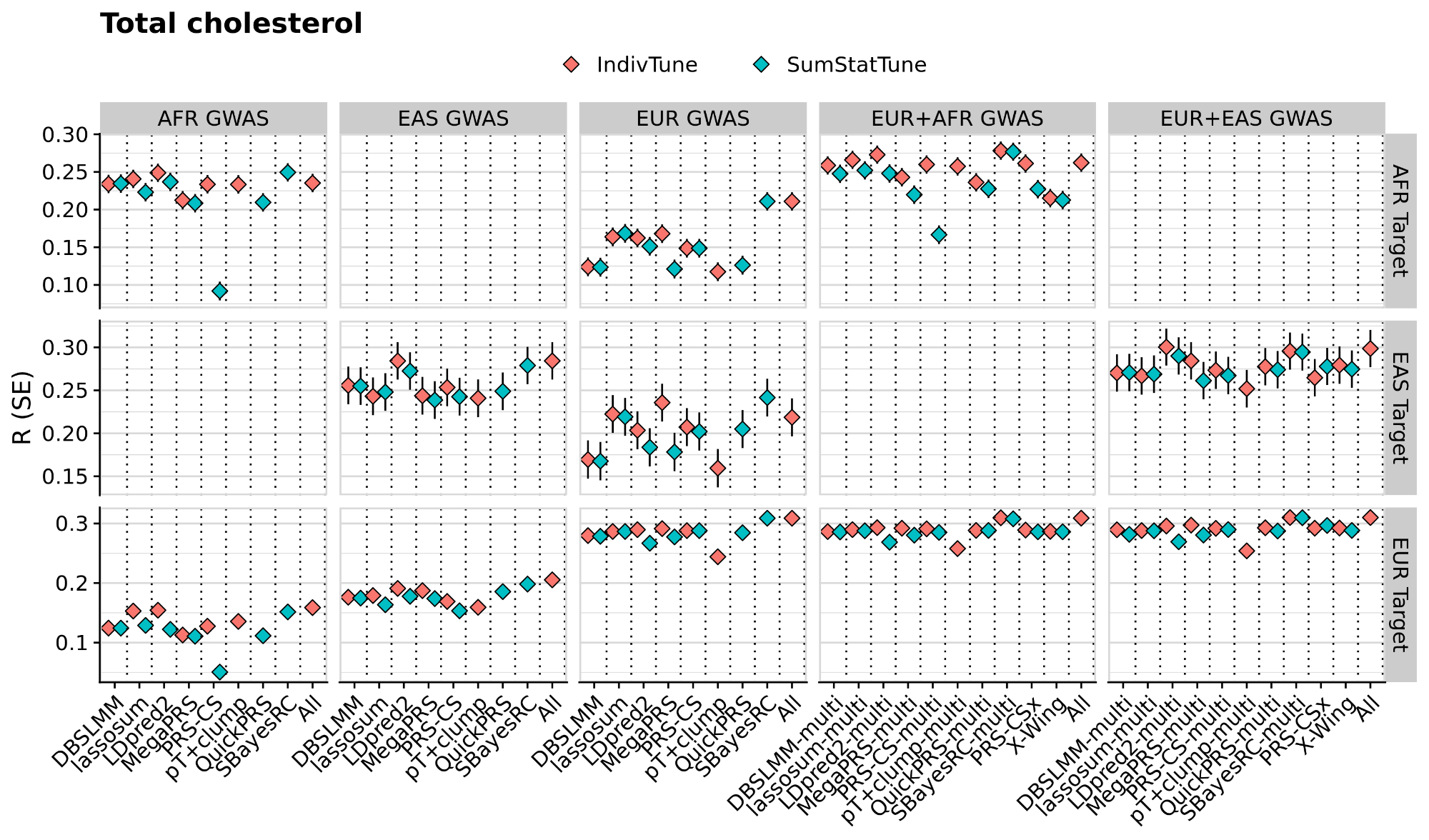

Figure S7. Predictive utility of PGS methods for Total cholesterol. The y-axis shows the correlation (R) between predicted and observed trait levels, with error bars representing the standard error. Colours differentiate between PGS methods trained using individual-level data (IndivTrain) and those trained using GWAS summary statistics (SumStatTrain). Facet columns indicate the GWAS data source used for PGS derivation, including African (AFR GWAS), East Asian (EAS GWAS), European (EUR GWAS), combined African and European (EUR+AFR GWAS), and combined East Asian and European (EUR+EAS GWAS) data. Facet rows represent performance in African (AFR Target), East Asian (EAS Target), and European (EUR Target) populations.

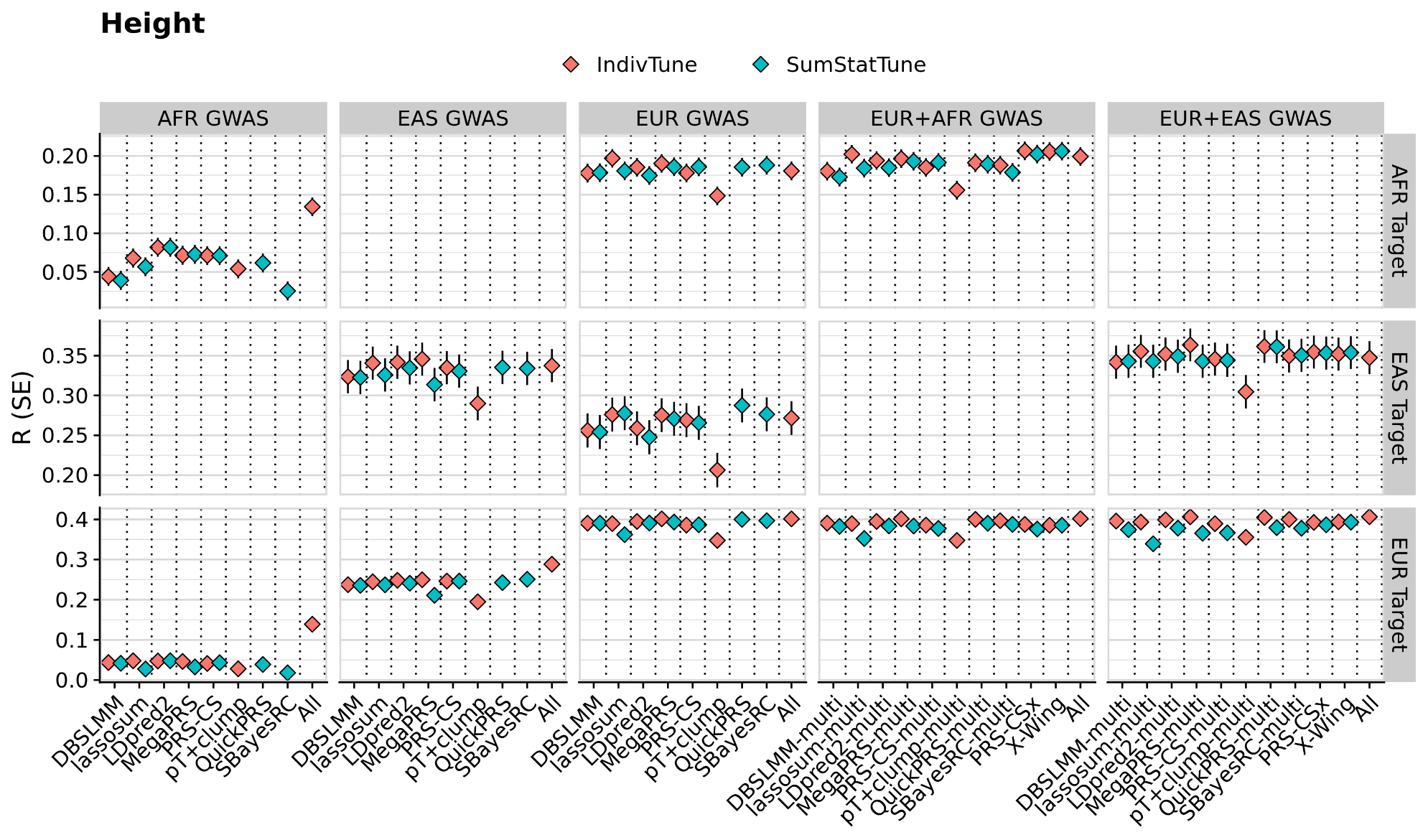

Figure S8. Predictive utility of PGS methods for Height. The y-axis shows the correlation (R) between predicted and observed trait levels, with error bars representing the standard error. Colours differentiate between PGS methods trained using individual-level data (IndivTrain) and those trained using GWAS summary statistics (SumStatTrain). Facet columns indicate the GWAS data source used for PGS derivation, including African (AFR GWAS), East Asian (EAS GWAS), European (EUR GWAS), combined African and European (EUR+AFR GWAS), and combined East Asian and European (EUR+EAS GWAS) data. Facet rows represent performance in African (AFR Target), East Asian (EAS Target), and European (EUR Target) populations.

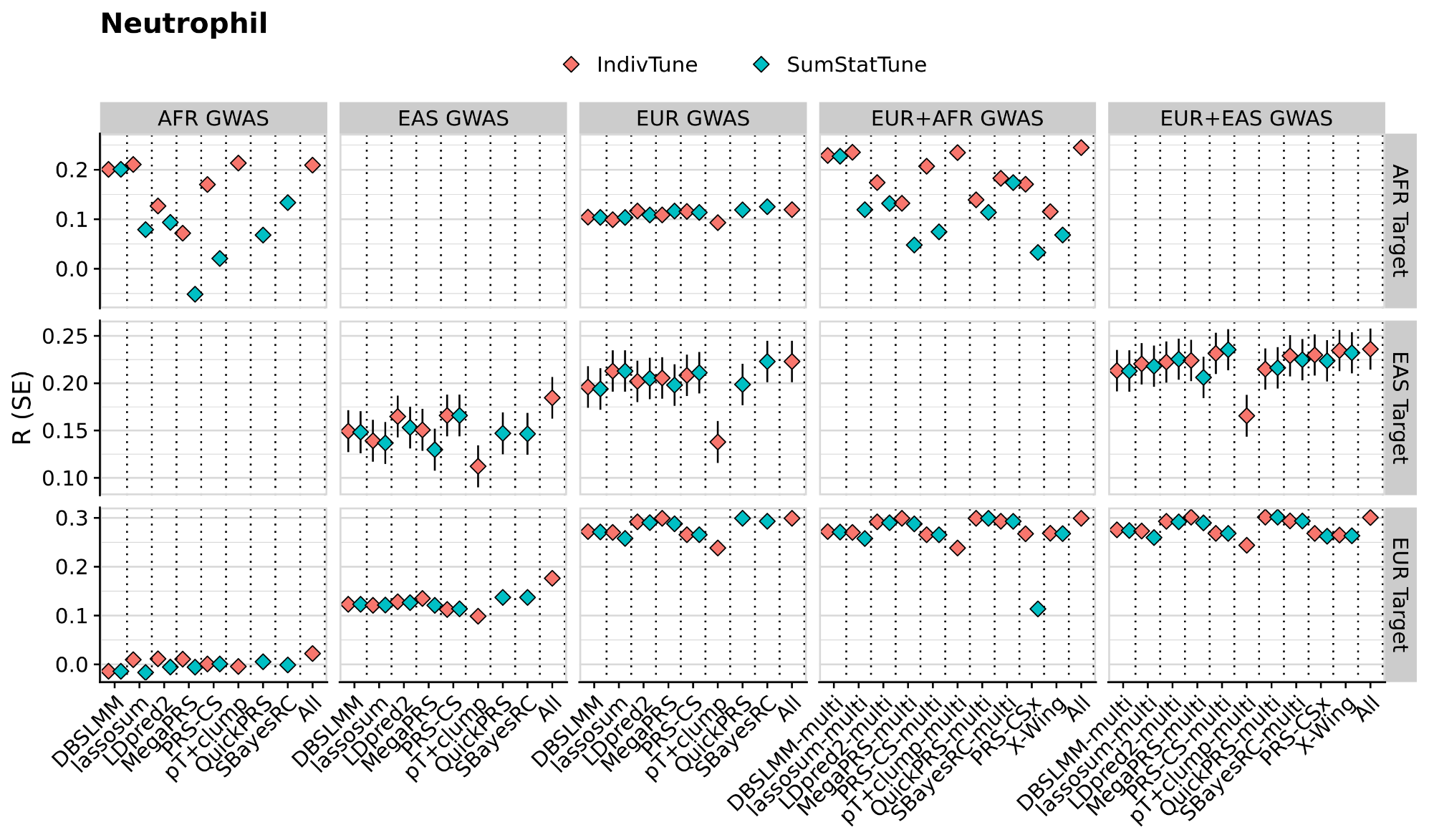

Figure S9. Predictive utility of PGS methods for Neutrophil. The y-axis shows the correlation (R) between predicted and observed trait levels, with error bars representing the standard error. Colours differentiate between PGS methods trained using individual-level data (IndivTrain) and those trained using GWAS summary statistics (SumStatTrain). Facet columns indicate the GWAS data source used for PGS derivation, including African (AFR GWAS), East Asian (EAS GWAS), European (EUR GWAS), combined African and European (EUR+AFR GWAS), and combined East Asian and European (EUR+EAS GWAS) data. Facet rows represent performance in African (AFR Target), East Asian (EAS Target), and European (EUR Target) populations.

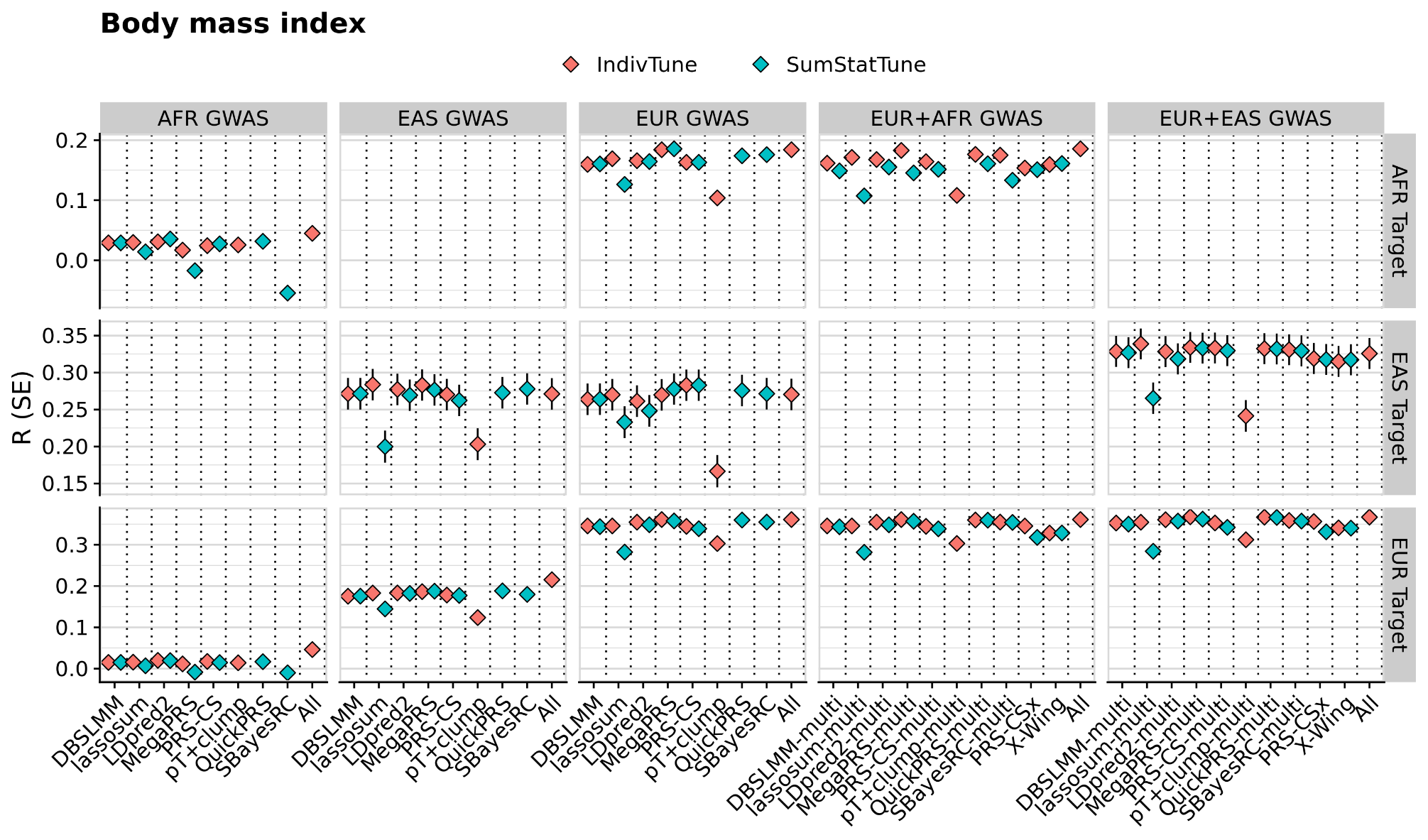

Figure S10. Predictive utility of PGS methods for Body mass index. The y-axis shows the correlation (R) between predicted and observed trait levels, with error bars representing the standard error. Colours differentiate between PGS methods trained using individual-level data (IndivTrain) and those trained using GWAS summary statistics (SumStatTrain). Facet columns indicate the GWAS data source used for PGS derivation, including African (AFR GWAS), East Asian (EAS GWAS), European (EUR GWAS), combined African and European (EUR+AFR GWAS), and combined East Asian and European (EUR+EAS GWAS) data. Facet rows represent performance in African (AFR Target), East Asian (EAS Target), and European (EUR Target) populations.

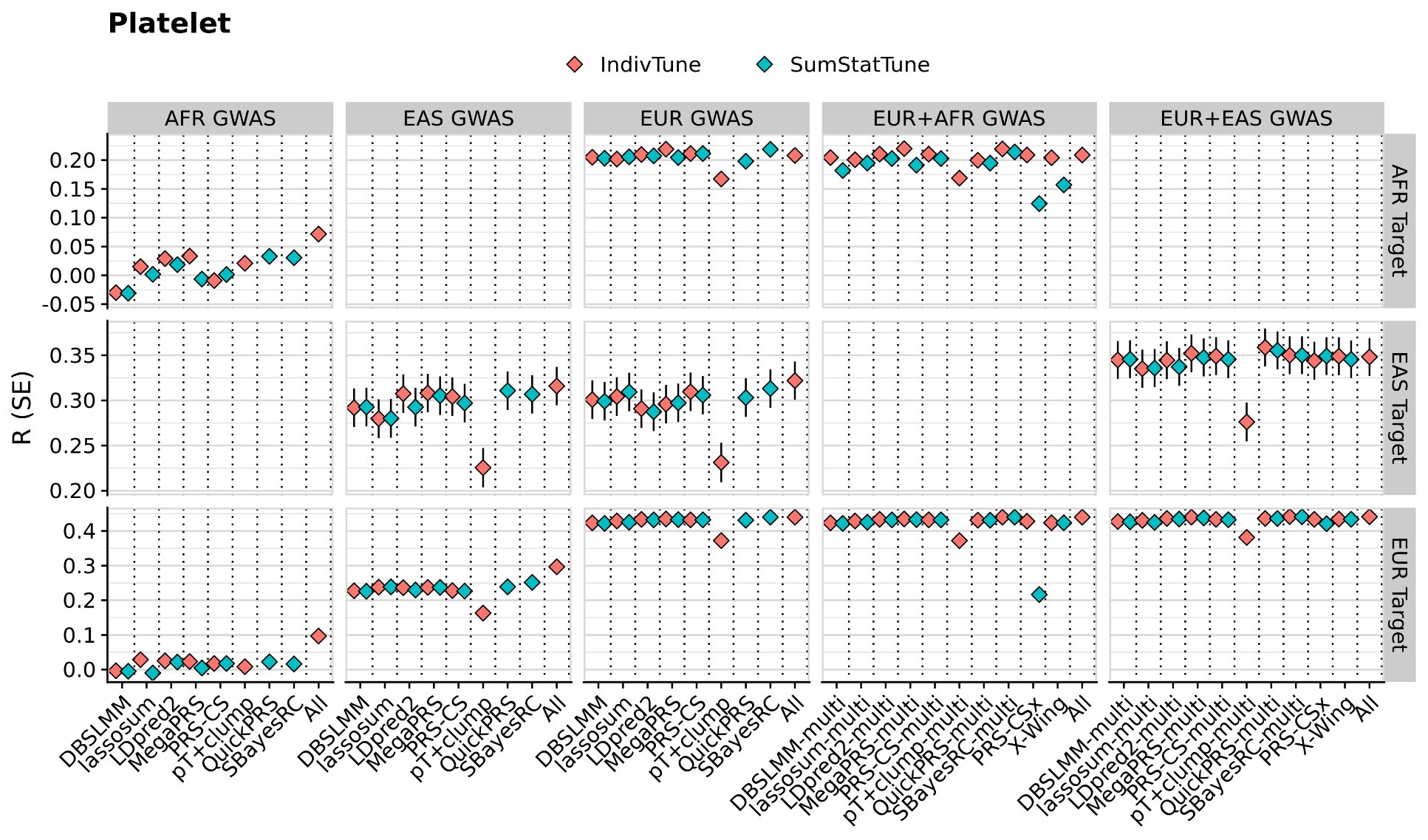

Figure S11. Predictive utility of PGS methods for Platelet. The y-axis shows the correlation (R) between predicted and observed trait levels, with error bars representing the standard error. Colours differentiate between PGS methods trained using individual-level data (IndivTrain) and those trained using GWAS summary statistics (SumStatTrain). Facet columns indicate the GWAS data source used for PGS derivation, including African (AFR GWAS), East Asian (EAS GWAS), European (EUR GWAS), combined African and European (EUR+AFR GWAS), and combined East Asian and European (EUR+EAS GWAS) data. Facet rows represent performance in African (AFR Target), East Asian (EAS Target), and European (EUR Target) populations.

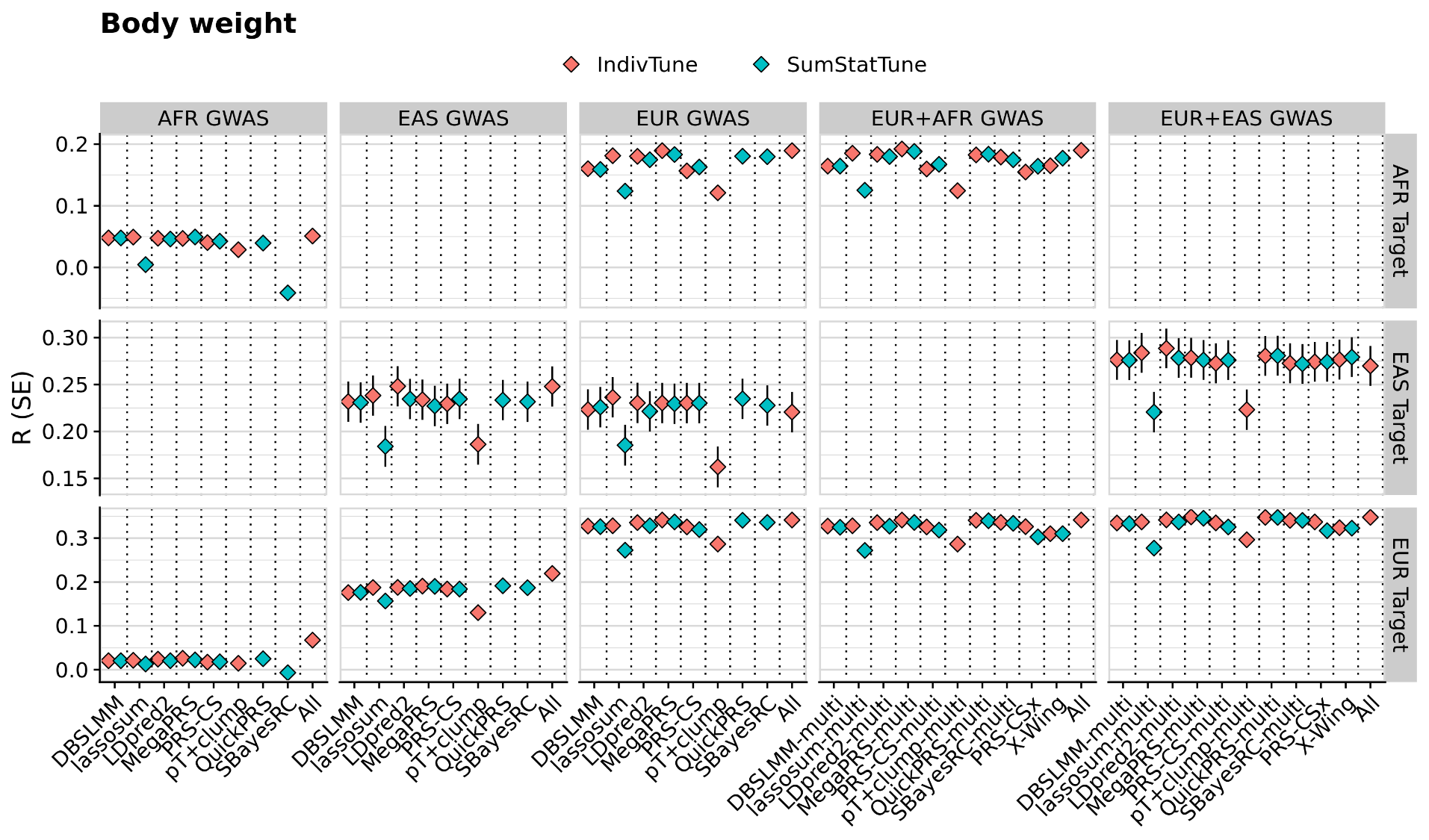

Figure S12. Predictive utility of PGS methods for Body weight. The y-axis shows the correlation (R) between predicted and observed trait levels, with error bars representing the standard error. Colours differentiate between PGS methods trained using individual-level data (IndivTrain) and those trained using GWAS summary statistics (SumStatTrain). Facet columns indicate the GWAS data source used for PGS derivation, including African (AFR GWAS), East Asian (EAS GWAS), European (EUR GWAS), combined African and European (EUR+AFR GWAS), and combined East Asian and European (EUR+EAS GWAS) data. Facet rows represent performance in African (AFR Target), East Asian (EAS Target), and European (EUR Target) populations.

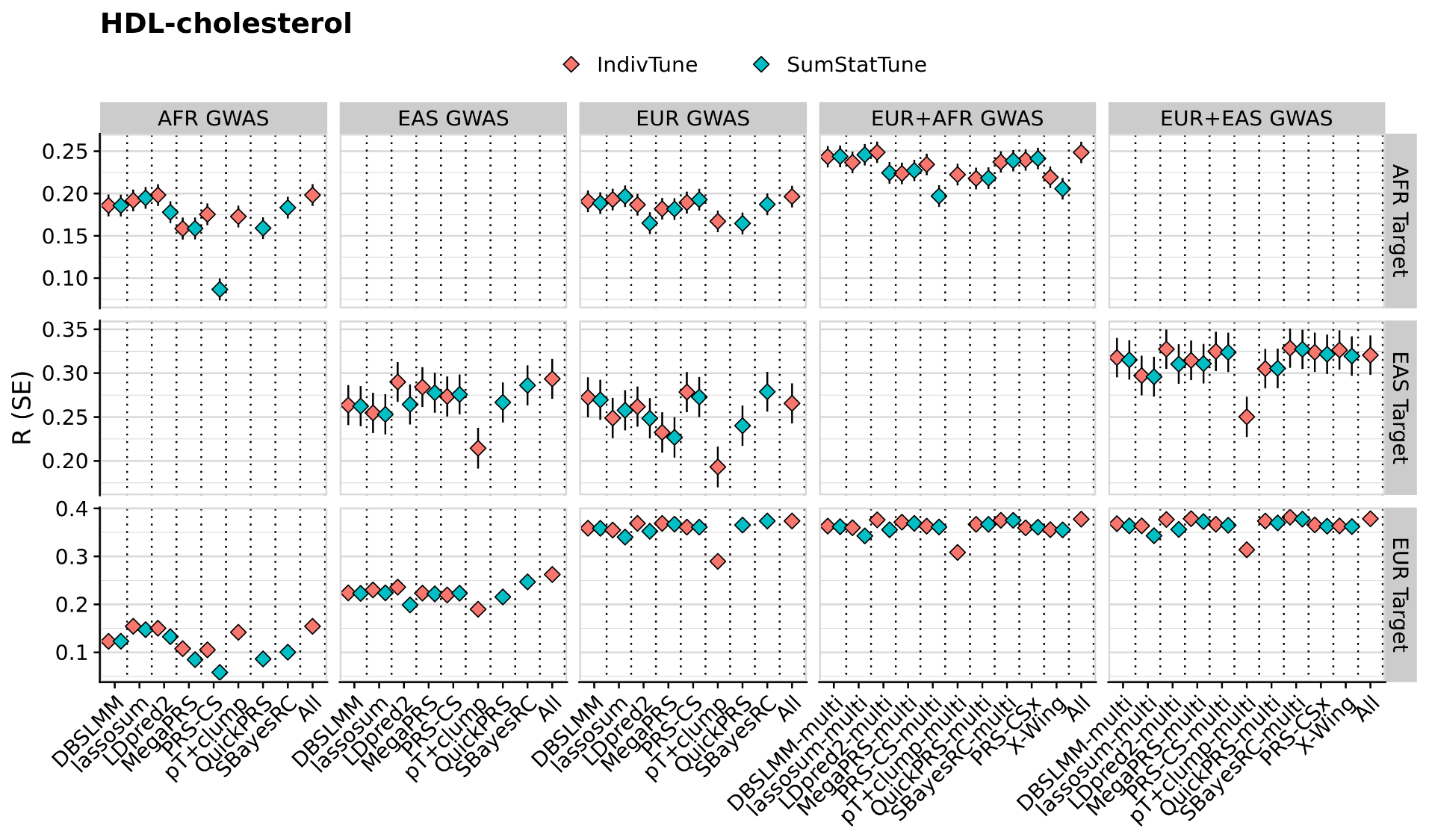

Figure S13. Predictive utility of PGS methods for HDL-cholesterol. The y-axis shows the correlation (R) between predicted and observed trait levels, with error bars representing the standard error. Colours differentiate between PGS methods trained using individual-level data (IndivTrain) and those trained using GWAS summary statistics (SumStatTrain). Facet columns indicate the GWAS data source used for PGS derivation, including African (AFR GWAS), East Asian (EAS GWAS), European (EUR GWAS), combined African and European (EUR+AFR GWAS), and combined East Asian and European (EUR+EAS GWAS) data. Facet rows represent performance in African (AFR Target), East Asian (EAS Target), and European (EUR Target) populations.

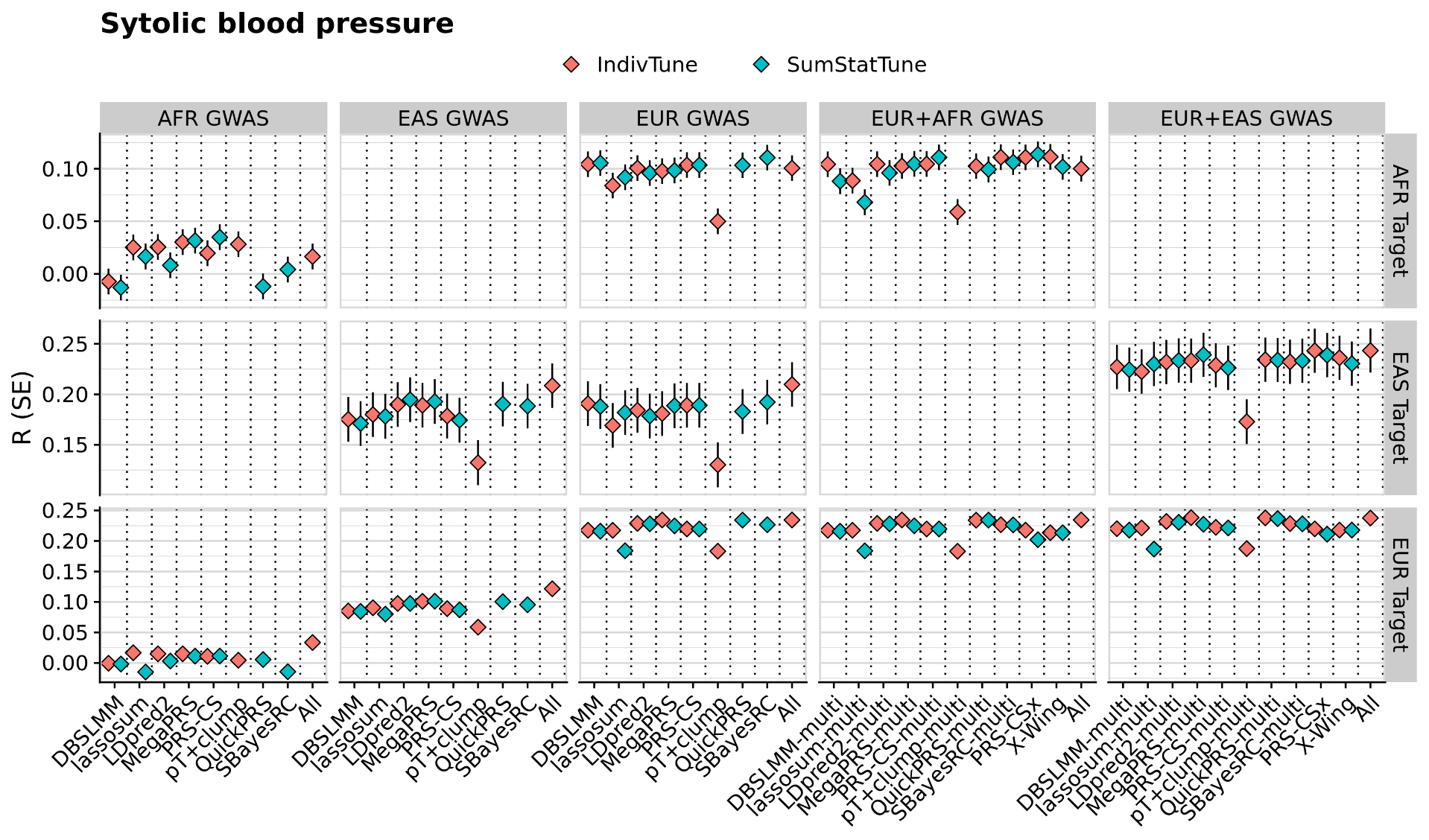

Figure S14. Predictive utility of PGS methods for Systolic blood pressure. The y-axis shows the correlation (R) between predicted and observed trait levels, with error bars representing the standard error. Colours differentiate between PGS methods trained using individual-level data (IndivTrain) and those trained using GWAS summary statistics (SumStatTrain). Facet columns indicate the GWAS data source used for PGS derivation, including African (AFR GWAS), East Asian (EAS GWAS), European (EUR GWAS), combined African and European (EUR+AFR GWAS), and combined East Asian and European (EUR+EAS GWAS) data. Facet rows represent performance in African (AFR Target), East Asian (EAS Target), and European (EUR Target) populations.

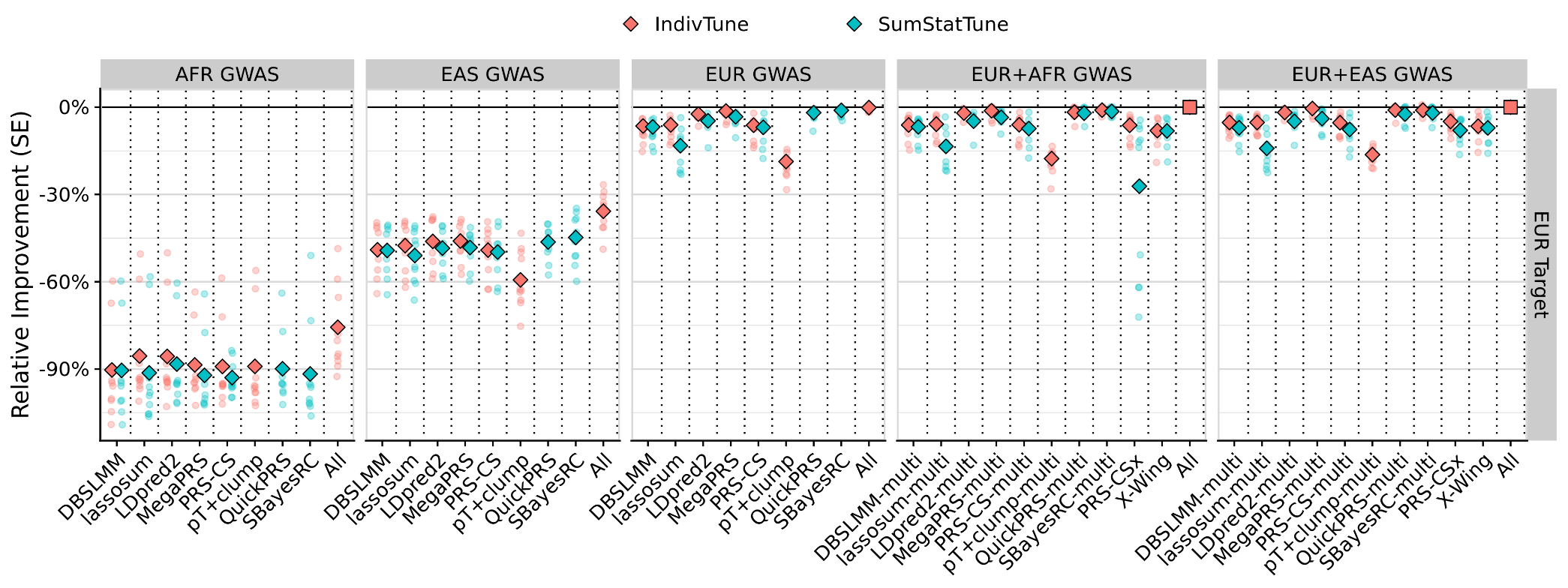

Figure S15. Relative predictive utility of PGS methods in the EUR target population. The y-axis shows the relative improvement in predictive performance compared to the multi-source ‘All’ model, with error bars representing the standard error. The diamond-shaped points indicate the average difference across traits, with small circular points indicating trait-specific differences. Colours indicate whether the PGS methods were trained using individual-level data (IndivTrain) or GWAS summary statistics (SumStatTrain). Facet columns represent the source of the GWAS data used for PGS derivation, including African (AFR GWAS), East Asian (EAS GWAS), European (EUR GWAS), combined African and European (EUR+AFR GWAS), and combined East Asian and European (EUR+EAS GWAS) data. This figure demonstrates the impact of different GWAS sources and training approaches on the predictive utility of PGS in the EUR target population.

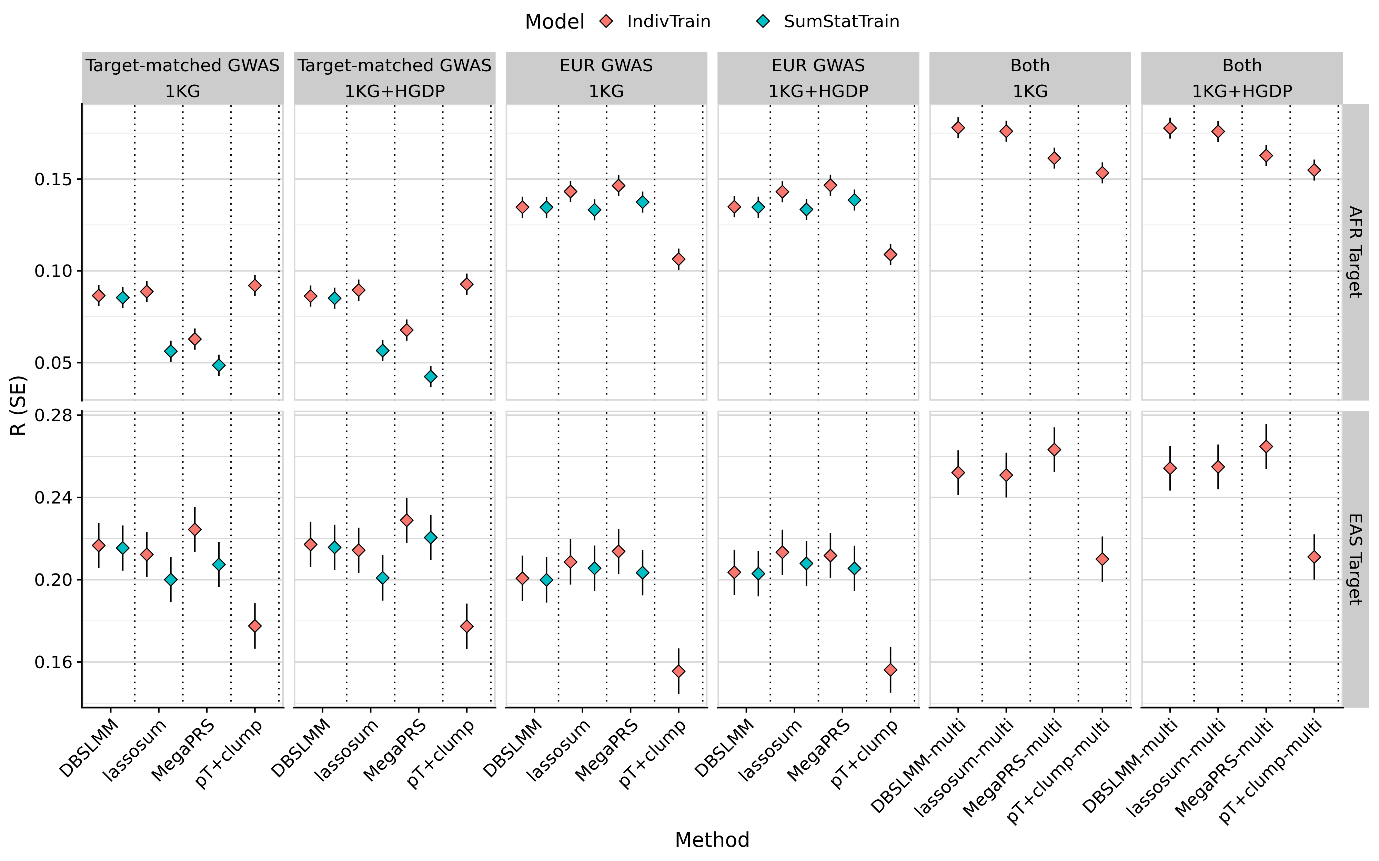

Figure S16. Comparing performance of PGS methods when using either 1KG or 1KG+HGDP reference data. The y-axis indicates the average correlation between predicted and observed values across traits, with error bars showing the standard error. Colours distinguish whether PGS methods were trained using individual-level data (IndivTrain) or GWAS summary statistics alone (SumStatTrain). 'Target-matched GWAS', 'EUR GWAS', and 'Both' facets show PGS performance using target ancestry-aligned, European, or combined GWAS data, respectively. ‘1KG’ and ‘1KG+HGDP’ facets show PGS performance using the 1KG or 1KG+HGDP as reference data. 'AFR Target' and 'EAS Target' facets show performance in AFR and EAS samples. There is no notable impact of the reference data on PGS performance for these methods.

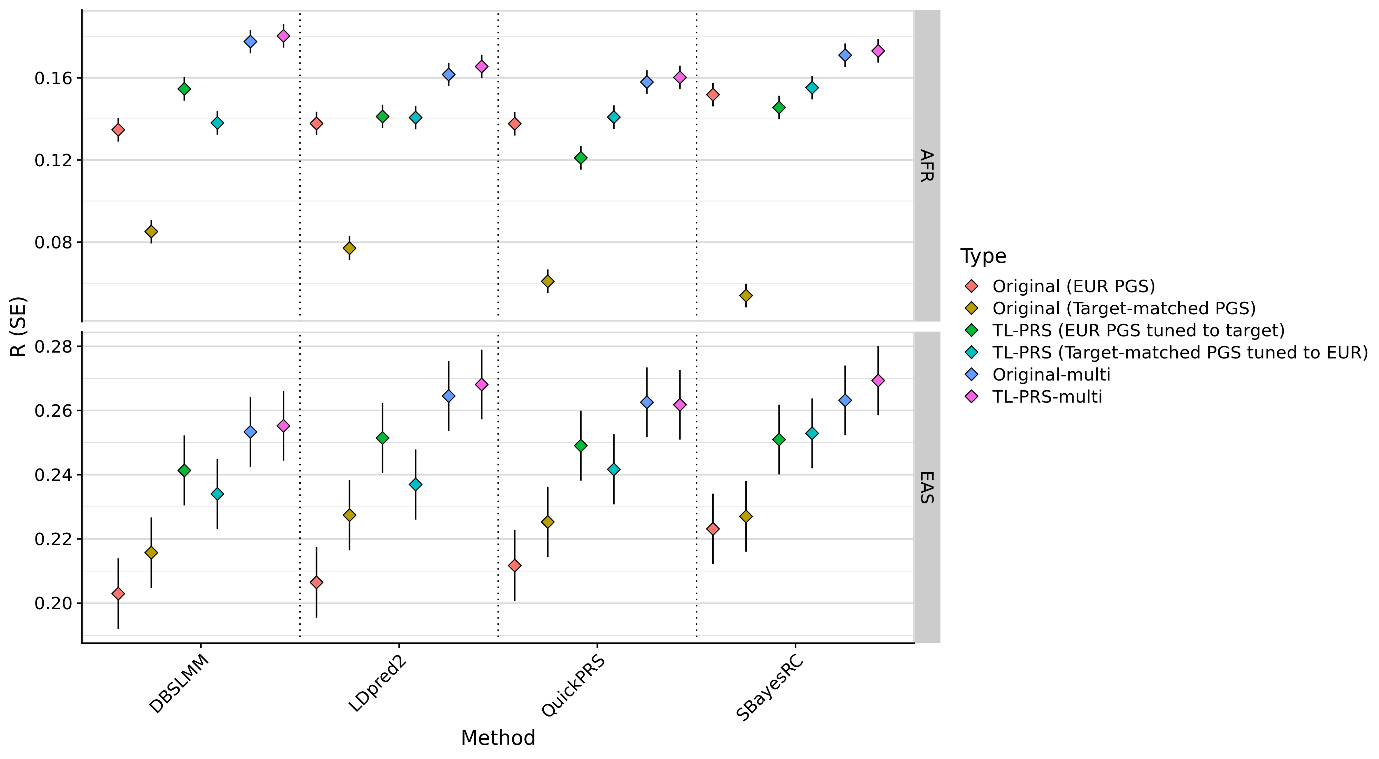

Figure S17. Comparing performance of TL-PRS adjusted PGS to unadjusted PGS. The y-axis indicates the average correlation between predicted and observed values across traits, with error bars showing the standard error. Colours distinguish different PGS models. ‘Original (EUR PGS)’ indicates a PGS derived using a EUR GWAS. ‘Original (Target-matched PGS)’ indicates a PGS derived using a target ancestry-aligned GWAS. ‘TL-PRS (EUR PGS tuned to target)’ indicates a EUR PGS that has been tuned towards the target population using TL-PRS. ‘TL-PRS (Target-matched PGS tuned to EUR)’ indicates a target ancestry-aligned PGS that has been tuned towards a EUR population using TL-PRS. ‘Original-multi’ indicates a model considering unadjusted PGS for both EUR and target-aligned populations, equivalent to the independently optimised multi-source approach. ‘TL-PRS-multi’ indicates a model considering TL-PRS-adjusted PGS for both EUR and target-aligned populations.

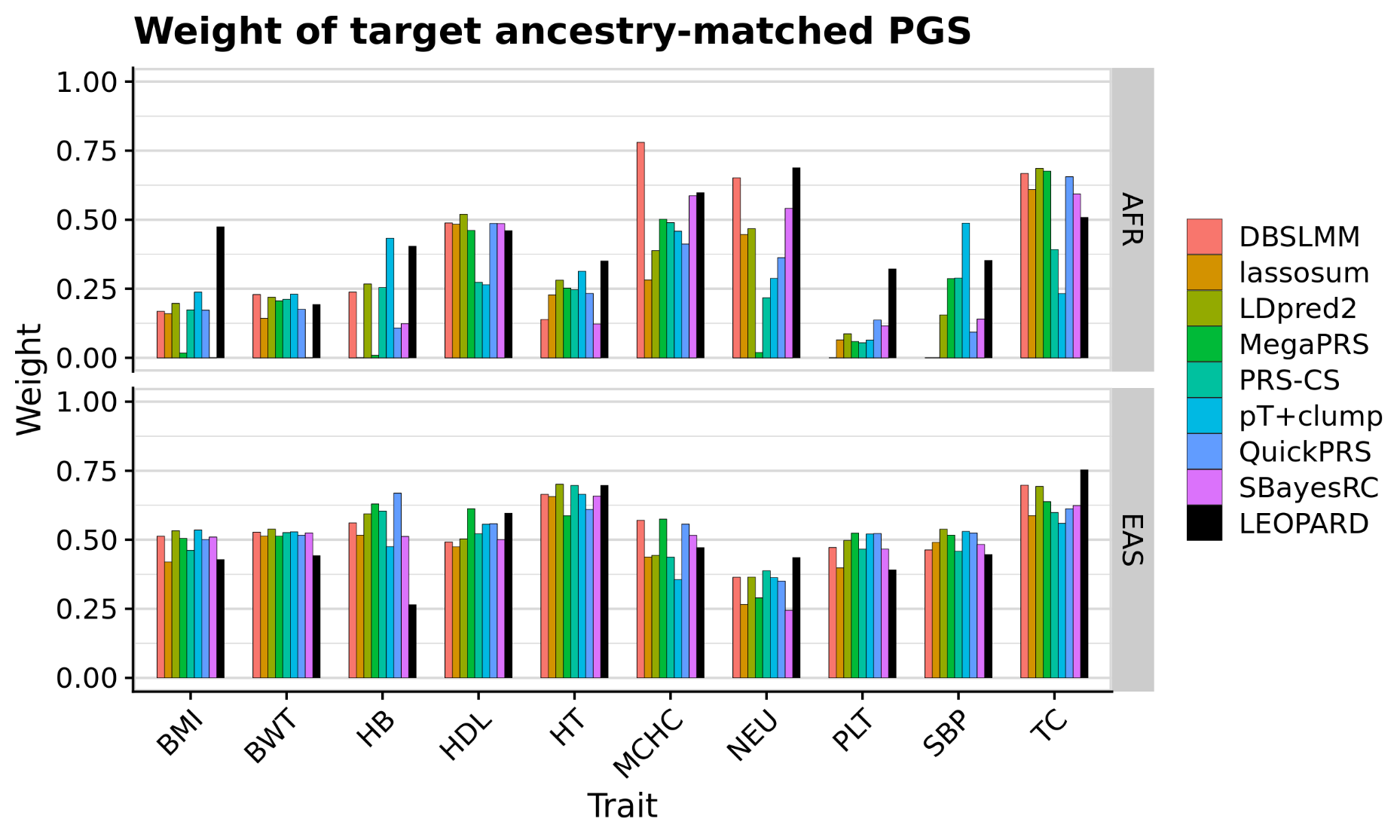

Figure S18. Observed weight of target ancestry-aligned PGS compared to weights estimated using LEOPARD (with QuickPRS). The y-axis represents the weight assigned to PGS for each method, with the x-axis showing different traits. Colours distinguish between PGS methods when estimating weights using individual-level data, with the black colour indicating weights estimated using LEOPARD (with QuickPRS). The top panel shows results for the African (AFR) target population, while the bottom panel shows results for the East Asian (EAS) target population. This figure highlights the differences in PGS weights derived using individual-level data compared to those estimated using the LEOPARD method when using AFR and EAS GWAS.

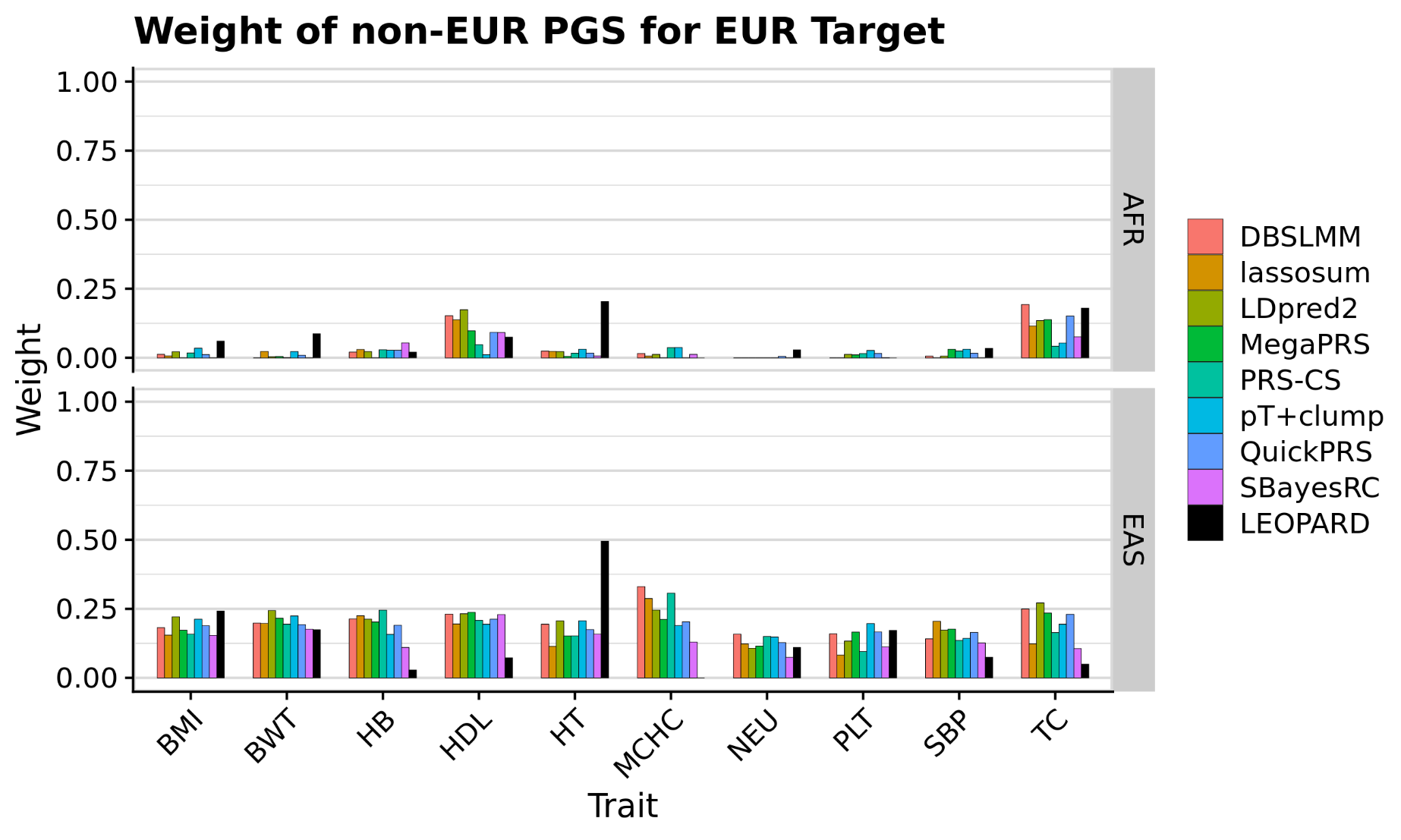

Figure S19. Observed weight of non-EUR PGS for EUR target population. The y-axis represents the weight assigned to polygenic scores (PGS) for each method, with the x-axis showing different traits. Colours differentiate between PGS methods when estimating weights using individual-level data, with black indicating weights estimated using the LEOPARD method (with QuickPRS). The top panel shows results using African (AFR) GWAS in combination with European (EUR) GWAS, while the bottom panel shows results using East Asian (EAS) GWAS in combination with EUR GWAS. This figure illustrates the relative contribution of non-EUR PGS when combined with EUR GWAS.

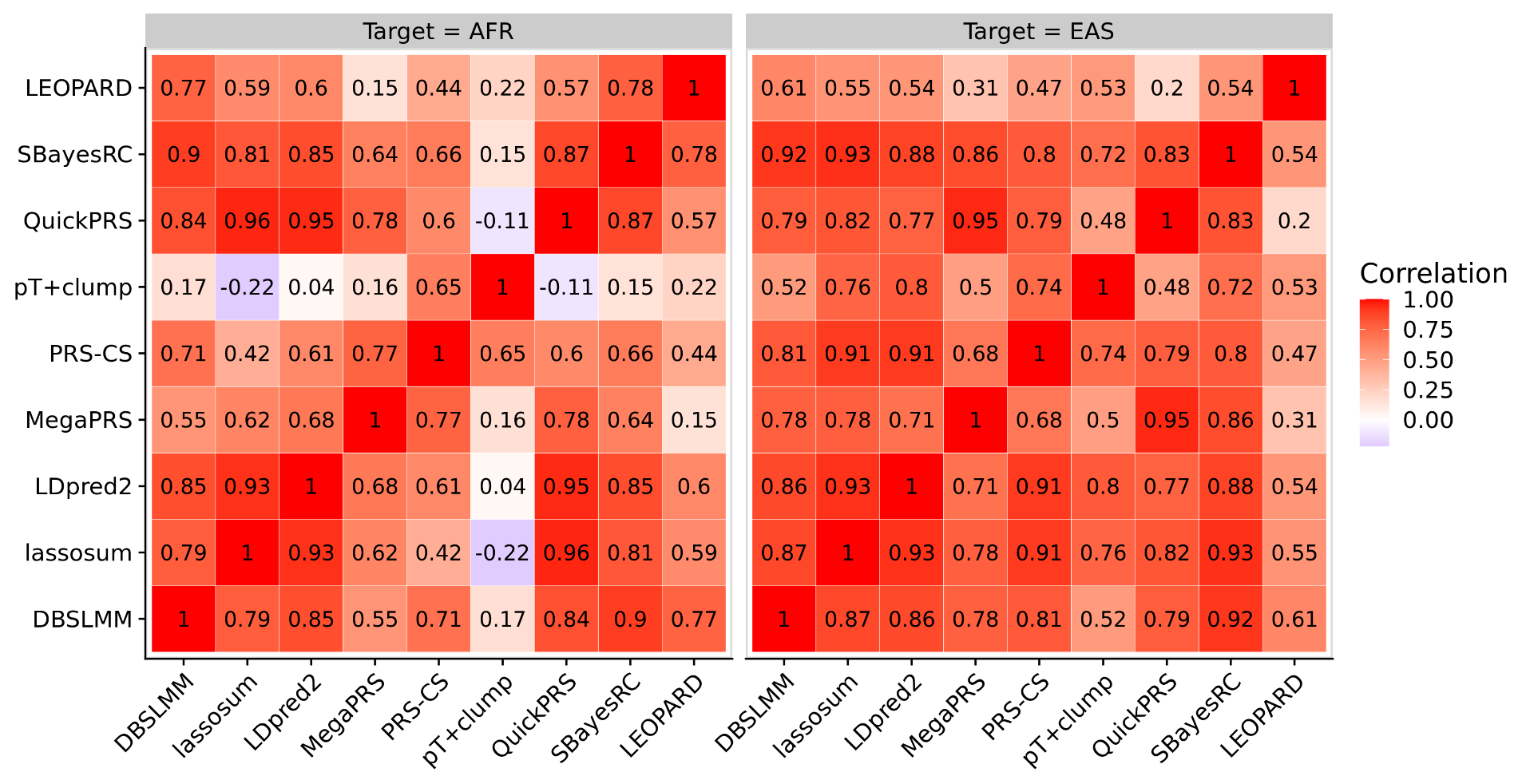

Figure S20. Correlation between observed PGS weights and LEOPARD-estimated weights for AFR and EAS target populations. The left panel shows results for the African (AFR) target population, while the right panel shows results for the East Asian (EAS) target population. Correlation values are represented by both colour intensity and numeric labels.

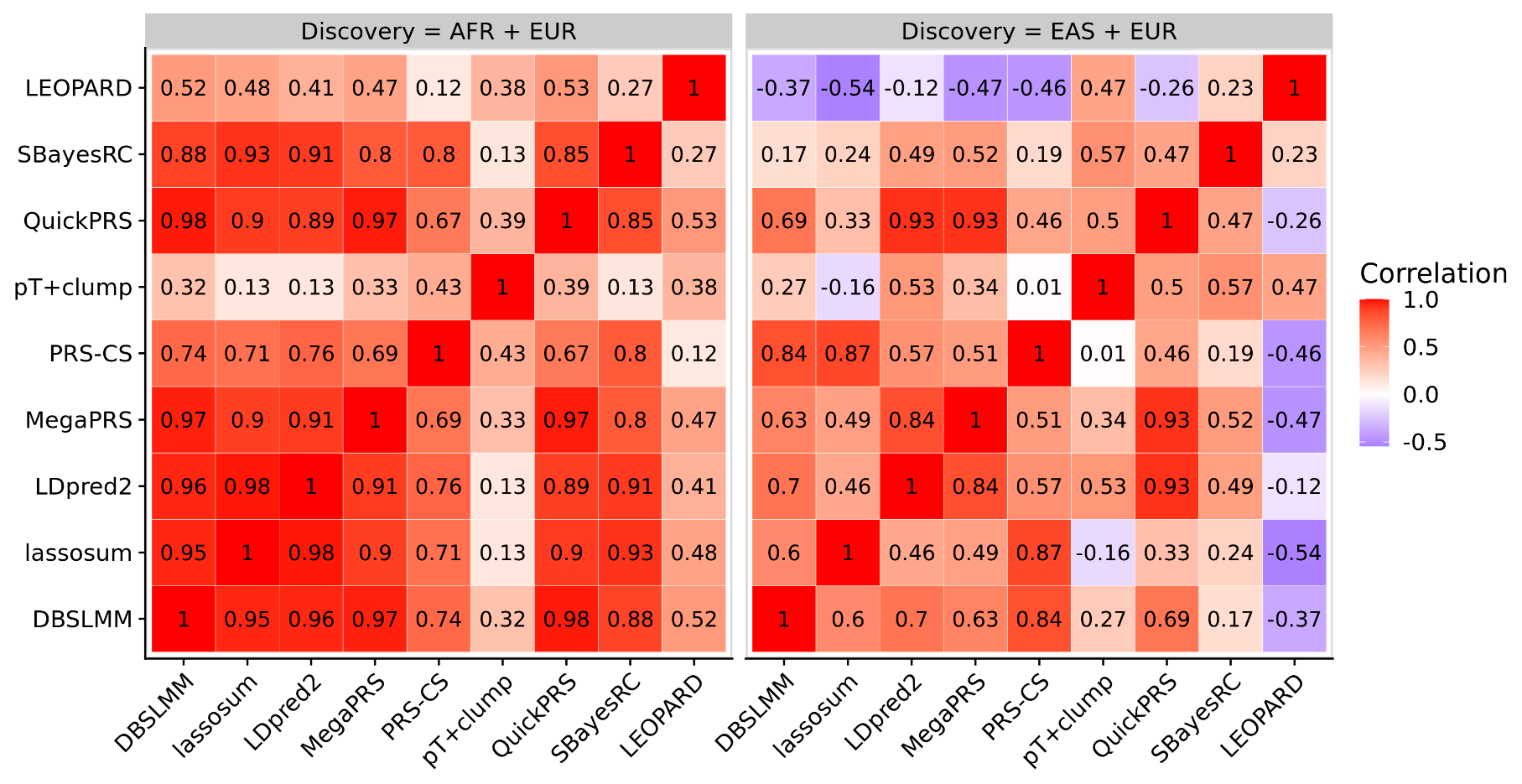

Figure S21. Correlation between observed PGS weights and LEOPARD-estimated weights in EUR samples. The left panel displays results for discovery datasets combining African (AFR) and European (EUR) GWAS, while the right panel shows results for combined East Asian (EAS) and EUR GWAS. Correlation values are represented both by colour intensity and numeric labels.

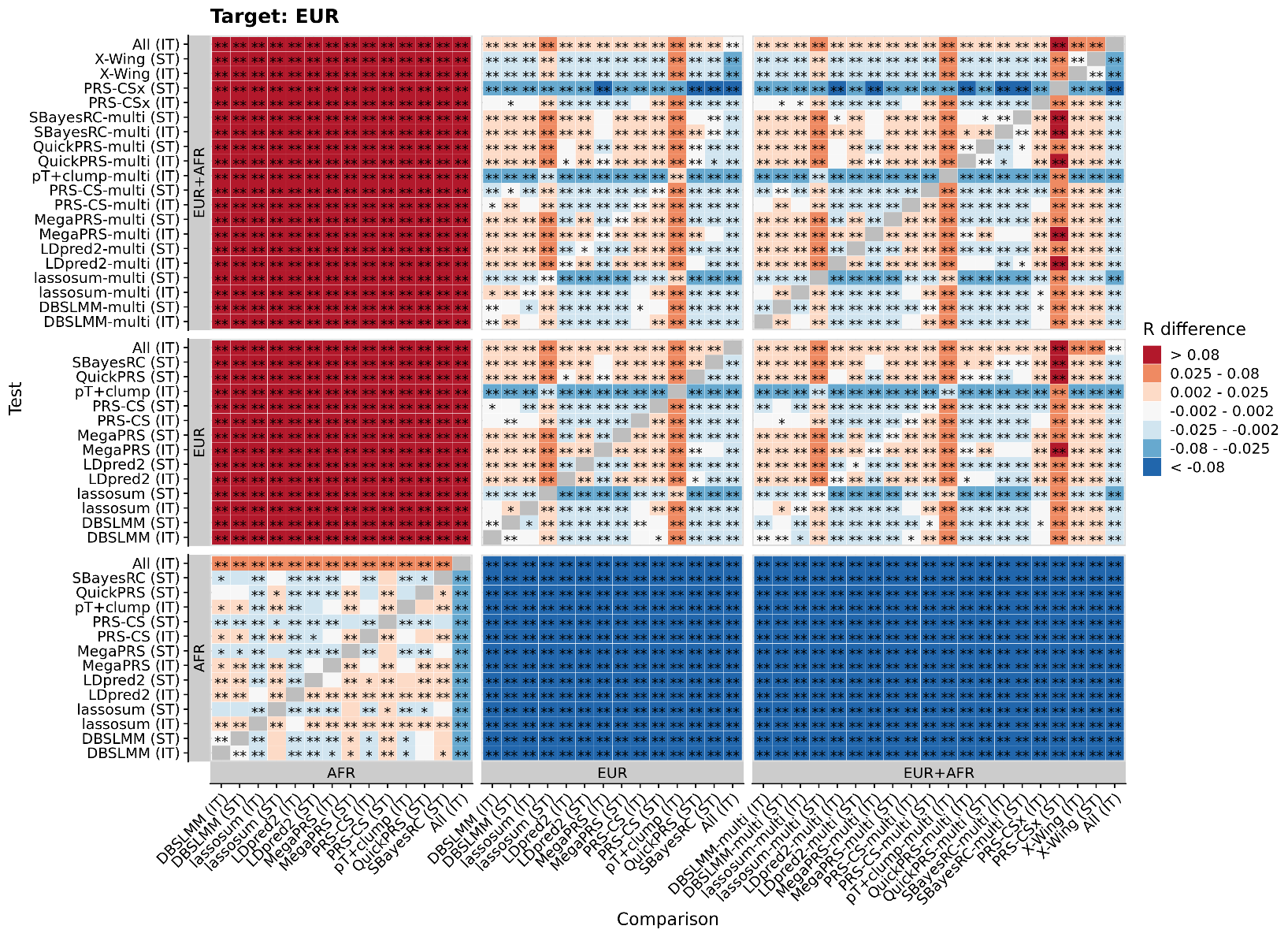

Figure S22. Pairwise comparison between all methods in EUR target sample, using EUR and AFR GWAS data, showing average difference in observed-expected correlation. R difference = Test correlation minus Comparison correlation. Red/orange colouring indicates the Test method (shown on Y axis) performed better than the Comparison method (shown on X axis). Shows only results based on the UKB target sample when using the 1KG reference. * = p<0.05 * = p<1×10−3. P-values are two-sided. IT = IndivTrain, PGS model tuned using individual-level data. ST = SumStatTrain, PGS model tuned using GWAS summary statistics alone.

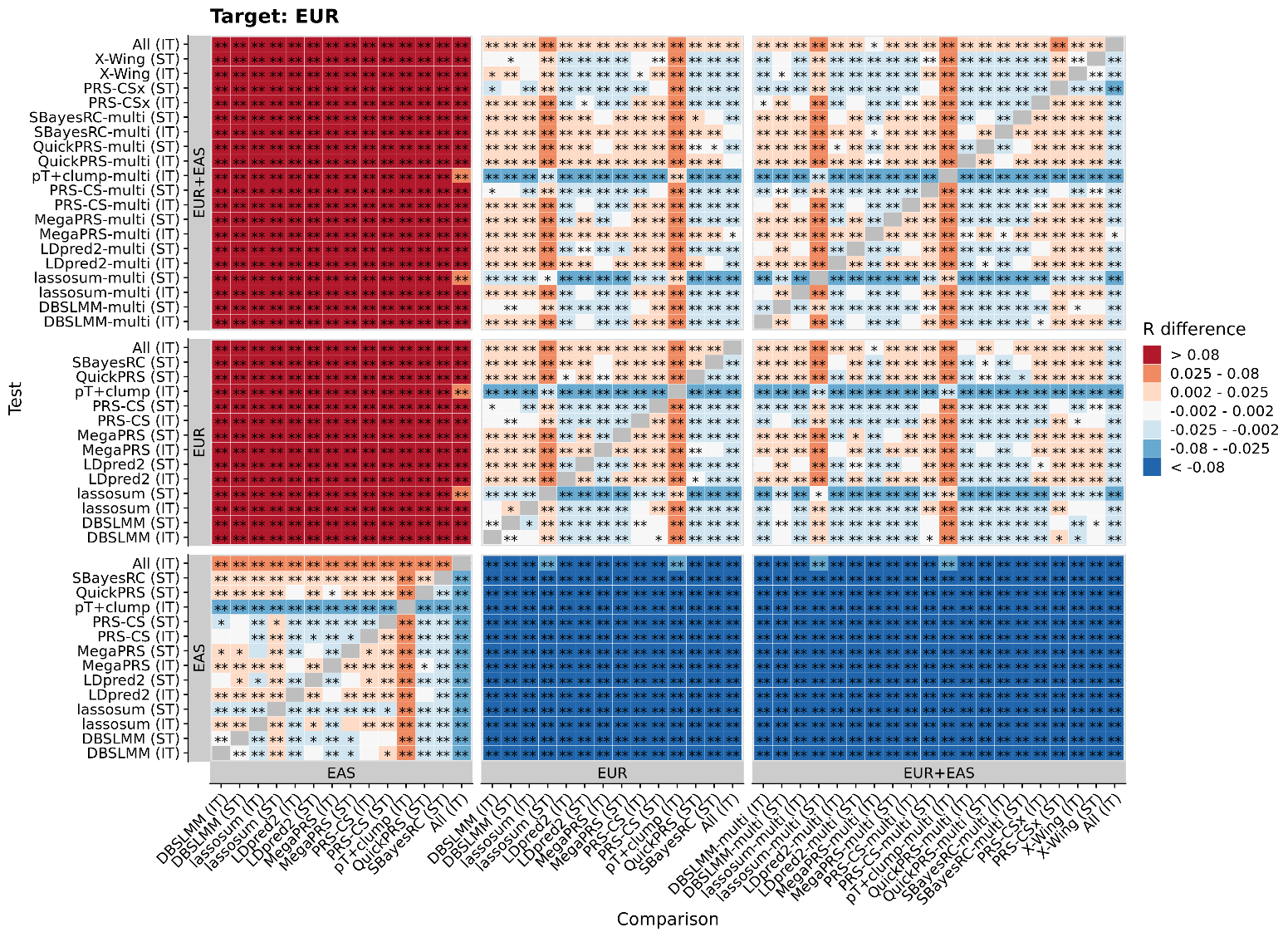

Figure S23. Pairwise comparison between all methods in EUR target sample, using EUR and EAS GWAS data, showing average difference in observed-expected correlation. R difference = Test correlation minus Comparison correlation. Red/orange colouring indicates the Test method (shown on Y axis) performed better than the Comparison method (shown on X axis). Shows only results based on the UKB target sample when using the 1KG reference. * = p<0.05 * = p<1×10−3. P-values are two-sided. IT = IndivTrain, PGS model tuned using individual-level data. ST = SumStatTrain, PGS model tuned using GWAS summary statistics alone.
